# Metformin reduces SARS-CoV-2 in a Phase 3 Randomized Placebo Controlled Clinical Trial

**DOI:** 10.1101/2023.06.06.23290989

**Authors:** Carolyn T Bramante, Kenneth B Beckman, Tanvi Mehta, Amy B Karger, David J Odde, Christopher J Tignanelli, John B Buse, Darrell M Johnson, Ray H. B. Watson, Jerry J Daniel, David M Liebovitz, Jacinda M Nicklas, Kenneth Cohen, Michael A Puskarich, Hrishikesh K Belani, Lianne K Siegel, Nichole R Klatt, Blake Anderson, Katrina M Hartman, Via Rao, Aubrey A Hagen, Barkha Patel, Sarah L Fenno, Nandini Avula, Neha V Reddy, Spencer M Erickson, Regina D. Fricton, Samuel Lee, Gwendolyn Griffiths, Matthew F Pullen, Jennifer L. Thompson, Nancy Sherwood, Thomas A. Murray, Michael R. Rose, David R Boulware, Jared D. Huling, the COVID-OUT study team

## Abstract

Current antiviral treatment options for SARS-CoV-2 infections are not available globally, cannot be used with many medications, and are limited to virus-specific targets.^1-3^ Biophysical modeling of SARS-CoV-2 replication predicted that protein translation is an especially attractive target for antiviral therapy.^4^ Literature review identified metformin, widely known as a treatment for diabetes, as a potential suppressor of protein translation via targeting of the host mTor pathway.^5^ In vitro, metformin has antiviral activity against RNA viruses including SARS-CoV-2.^6,7^ In the COVID-OUT phase 3, randomized, placebo-controlled trial of outpatient treatment of COVID-19, metformin had a 42% reduction in ER visits/hospitalizations/death through 14 days; a 58% reduction in hospitalizations/death through 28 days, and a 42% reduction in Long COVID through 10 months.^8,9^ Here we show viral load analysis of specimens collected in the COVID-OUT trial that the mean SARS-CoV-2 viral load was reduced 3.6-fold with metformin relative to placebo (−0.56 log_10_ copies/mL; 95%CI, -1.05 to -0.06, p=0.027) while there was no virologic effect for ivermectin or fluvoxamine vs placebo. The metformin effect was consistent across subgroups and with emerging data.^10,11^ Our results demonstrate, consistent with model predictions, that a safe, widely available,^12^ well-tolerated, and inexpensive oral medication, metformin, can be repurposed to significantly reduce SARS-CoV-2 viral load.

## Main

COVID-OUT was a multi-site, decentralized, phase 3, quadruple-blinded, placebo controlled randomized clinical trial (RCT) testing outpatient treatment of coronavirus 2019 disease (COVID-19). ^1^ The clinical outcomes showed that immediate release metformin reduced the relative risk of emergency department visits / hospitalization / and death through 14-days by 42%; the relative risk of hospitalization or death through 28-days by 58%; and the relative risk of Long COVID through 10 months by over 40%.^1,2^

The COVID-OUT trial was motivated by *in silico* modeling, *in vitro* cell culture, and human lung tissue data that metformin decreased growth of severe acute respiratory coronavirus 2 (SARS-CoV-2) and improved cell viability. ^3-5^ The *in silico* modeling identified protein translation as a key process in SARS-CoV-2 replication.^3^ Metformin inhibits the mechanistic target of rapamycin (mTOR), ^6^ which controls protein translation and some of the first papers published on metformin describe actions against influenza, parainfluenza, and other viruses. ^7,8^ More recently, metformin was observed to have *in vitro* antiviral actions against Zika and hepatitis C, both RNA viruses. ^9-12^

Here we present the results of viral load quantification obtained during the COVID-OUT clinical trial. The trial was remotely delivered and used a 2 by 3 factorial design trial of parallel treatment arms to allow efficient assessment of three oral, generic medications: immediate release metformin; ivermectin; and fluvoxamine. The doses used for each medication had not been previously studied in a phase 3 clinical trial of outpatient treatment for COVID-19. Participants self-collected viral load samples from the anterior nares on Day 1, 5, and 10 as an optional component of this clinical trial.

## Methods

### Study Design, Sample, and Oversight

COVID-OUT was an investigator-initiated, multi-site, phase 3, randomized, quadruple-blinded placebo-controlled clinical trial (ClinicalTrials.gov: NCT04510194). ^1^ The trial enrolled from Dec 30, 2020 to Jan 28, 2022 and was decentralized to prevent in-person contact and potential spread of SARS-CoV-2. The participants, care providers, investigators, and outcomes assessors, including all laboratory personnel processing samples, are blinded to treatment allocation.

Protocol approval was provided by institutional review boards (IRBs) at each site, the Advarra central IRB. An independent data safety monitoring board (DSMB) monitored safety and efficacy. An independent monitor oversaw study conduct per the Declaration of Helsinki, Good Clinical Practice Guidelines, and local requirements.

COVID-OUT excluded low-risk individuals, limiting enrollment to average risk adults aged 30 to 85 years with a body mass index (BMI) in the overweight category or higher; documented positive SARS-CoV-2 within 3 days; no prior confirmed SARS-CoV-2 infection. Exclusion criteria: hospitalized; symptom onset >7 days prior; unstable heart, liver, or kidney failure. Additional detail may be found online. ^1^

Metformin was administered as 500mg on Day 1, 500mg twice daily on Days 2-5 and 500mg in the AM and 1,000mg in the PM on Days 6-14. Fluvoxamine was administered as 50mg on Day 1 and 50mg twice daily on Days 2-14. Ivermectin was administered at a median dose of 430mcg/kg/day (range 390 to 470mcg/kg/day) for three days.

### Clinical and Virologic Endpoints

The primary endpoint of the trial was severe COVID-19 by Day 14, defined as a binary, composite endpoint with 4 components: hypoxemia on home oximeter, emergency department visit, hospitalization, or death due to COVID-19. Secondary endpoints included clinical progression to hospitalization or death by Day 28 and Long COVID over 10-month follow-up. The virologic secondary endpoint was change in viral load from baseline to follow-up.

Self-collection of anterior nares samples was an optional component of the randomized trial. All participants were sent the materials for self-collecting nasal swabs. Supply chain shortages caused administrative censoring of 78 participants who did not receive materials for collecting Day 1, 5, or 10 samples; and 3 did not receive materials for collecting Day 5 or 10 samples **(Figure S1)**.

### Laboratory procedures

Participants received written instructions with pictures on collecting a nares swab sample from the anterior mid-turbinate. Viral load was measured via RT-qPCR using N1 and N2 targets in the SARS-CoV-2 nucleocapsid protein, with relative cycle threshold (Ct) values converted to absolute copy number via calibration to droplet digital PCR. Detailed methods can be found in **Table S10**.

While participant self-collection may vary between participants, self-collection of samples is done by the same individual at baseline and follow-up. Thus, participant self-collection may have less variability between baseline and follow-up than when study or clinical staff obtain samples.

### Statistical analysis

We evaluated study drug impact on log_10_-transformed viral load on day 5 and day 10 with a linear Tobit regression model where the effect of study drugs was allowed to differ on day 5 and day 10. This was decided a priori as a rigorous analytic approach to account for left censoring due to the viral load limit of quantification. Repeated measures were accounted for using clustered standard errors within participants. Analyses of viral loads estimated the adjusted mean reduction averaged over time and the adjusted mean reduction at day 5 and at day 10. We evaluated impact over time on the probability of viral load being undetectable using generalized estimating equations with a logistic link; estimates are reported as odds ratios and risk differences.

The COVID-OUT trial was a 2 by 3 factorial design of parallel distinct treatments (**Table S2)**. All analyses were adjusted for baseline viral load, vaccination status, time since last vaccination for those vaccinated before enrollment, receipt of other study medications within factorial trial, lab that processed the nasal swabs, and exact time/date of specimen collection. Additional detail and the results of the analysis with dropping of adjustment variables are presented in **Table S8, Table S9**.

To handle missing values, we used multiple imputation with chained equations to multiply impute missing viral load outcomes and vaccination status. Missing covariate information was jointly imputed along with missing outcomes using random forests for the univariate imputation models; along with outcome and vaccination status information, imputation models were informed by sex, BMI, symptom duration, race/ethnicity, baseline comorbidities, clinical outcomes, and enrollment time categorized by the dominant pandemic variant. Complete case analysis without imputation of missing data is presented in **Figures S2-S4**.

## Results

Among 1323 randomized participants in the COVID-OUT trial, 999 (76%) chose to participate in the optional sub-study and provided at least one self-administered nasal swab sample (**Table 1, Figure S1**). The demographics of the participants submitting swabs were similar to those who did not submit any nasal swabs (**Table S3**). Day 1 samples were provided by 945 participants; 871 provided Day 5 samples; and 775 provided Day 10 samples (**Table S6)**. The overall viral load was a median of 4.88 log_10_copies/mL (IQR, 2.99 to 6.18), on Day 1; 1.90 (IQR, 0 to 3.93) on Day 5, and 0 (IQR 0 to 1.90 with 0 representing the limit of quantification) on Day 10.

**Table 1.** Baseline characteristics of participants who submitted any nasal swab.

| Variable |  | Overall<br>N = 999 | Placebo<br>N = 495 | Metformin<br>N = 504 |
| --- | --- | --- | --- | --- |
| Age |  | 46 (38, 55) | 45 (38, 54) | 46 (38, 55) |
| Biologic Sex, Female |  | 56% (559) | 57% (282) | 55% (277) |
| Race | Native American | 2.2% (22) | 2.6% (13) | 1.8% (9) |
|  | Asian | 3.6% (36) | 3.8% (19) | 3.4% (17) |
|  | Hawaiian, Pacific Islander | 0.7% (7) | 0.4% (2) | 1.0% (5) |
|  | Black or African American | 6.2% (62) | 6.1% (30) | 6.3% (32) |
|  | White | 85% (849) | 85% (420) | 85% (429) |
|  | Other, missing, declined | 5.0% (50) | 4.4% (22) | 5.6% (28) |
| Ethnicity | Hispanic | 12% (118) | 13% (63) | 11% (55) |
| **Medical history** |  |  |  |  |
| BMI |  | 30.0 (27.1, 34.3) | 30.0 (26.9, 34.7) | 29.8 (27.2, 34.0) |
| BMI >= 30kg/m2 |  | 50% (496) | 51% (250) | 49% (246) |
| Cardiovascular disease |  | 28% (282) | 28% (140) | 28% (142) |
| Diabetes |  | 2.0% (20) | 2.6% (13) | 1.4% (7) |
| **Vaccination Status at baseline** |  |  |  |  |
| No Vaccine |  | 46% (457) | 48% (240) | 43% (217) |
| Primary Series Only |  | 50% (495) | 47% (232) | 52% (263) |
| Monovalent booster |  | 4.7% (47) | 4.6% (23) | 4.8% (24) |
| Days since last vaccine dose |  | 194 (132, 240) | 195 (132, 235) | 192 (132, 245) |
| **Time from symptom onset to first dose\*** |  |  |  |  |
| Days, mean (± SD) | | 4.7 (±1.9) | 4.7 (±1.8) | 4.7 (±1.9) |
| <= 4 Days |  | 46% (453) | 48% (230) | 45% (223) |
| **SARS-CoV-2 Variant Period** |  |  |  |  |
| Alpha (pre- June 19, 2021) |  | 13% (132) | 13% (65) | 13% (67) |
| Delta (June 19, 2021 to Dec 12, 2021) |  | 65% (645) | 65% (320) | 64% (325) |
| Omicron (after Dec 12, 2021) |  | 22% (222) | 22% (110) | 22% (112) |
| **Insurance status** |  |  |  |  |
| Private |  | 65% (652) | 65% (324) | 65% (328) |
| Medicare |  | 7.5% (75) | 6.9% (34) | 8.1% (41) |
| Medicaid |  | 14% (136) | 14% (69) | 13% (67) |
| No insurance |  | 12% (123) | 12% (60) | 12% (63) |
| Unknown |  | 1.3% (13) | 1.6% (8) | 1.0% (5) |

The overall mean SARS-CoV-2 viral load reduction with metformin was -0.56 log_10_ copies/mL (95%CI, -1.05 to -0.06) greater than placebo across all follow-up, p=0.027. The antiviral effect of metformin compared to placebo was -0.47 log_10_ copies/mL (95% CI -1.14 to 0.26) on Day 5 and -0.67 log_10_ copies/mL (95%CI, -1.80 to 0.44) on Day 10, **Figure 1**. Neither ivermectin nor fluvoxamine had any virologic effect at Day 5 or 10 (**Figures 2 and S2, Table S7)**.

**Figure 1.**
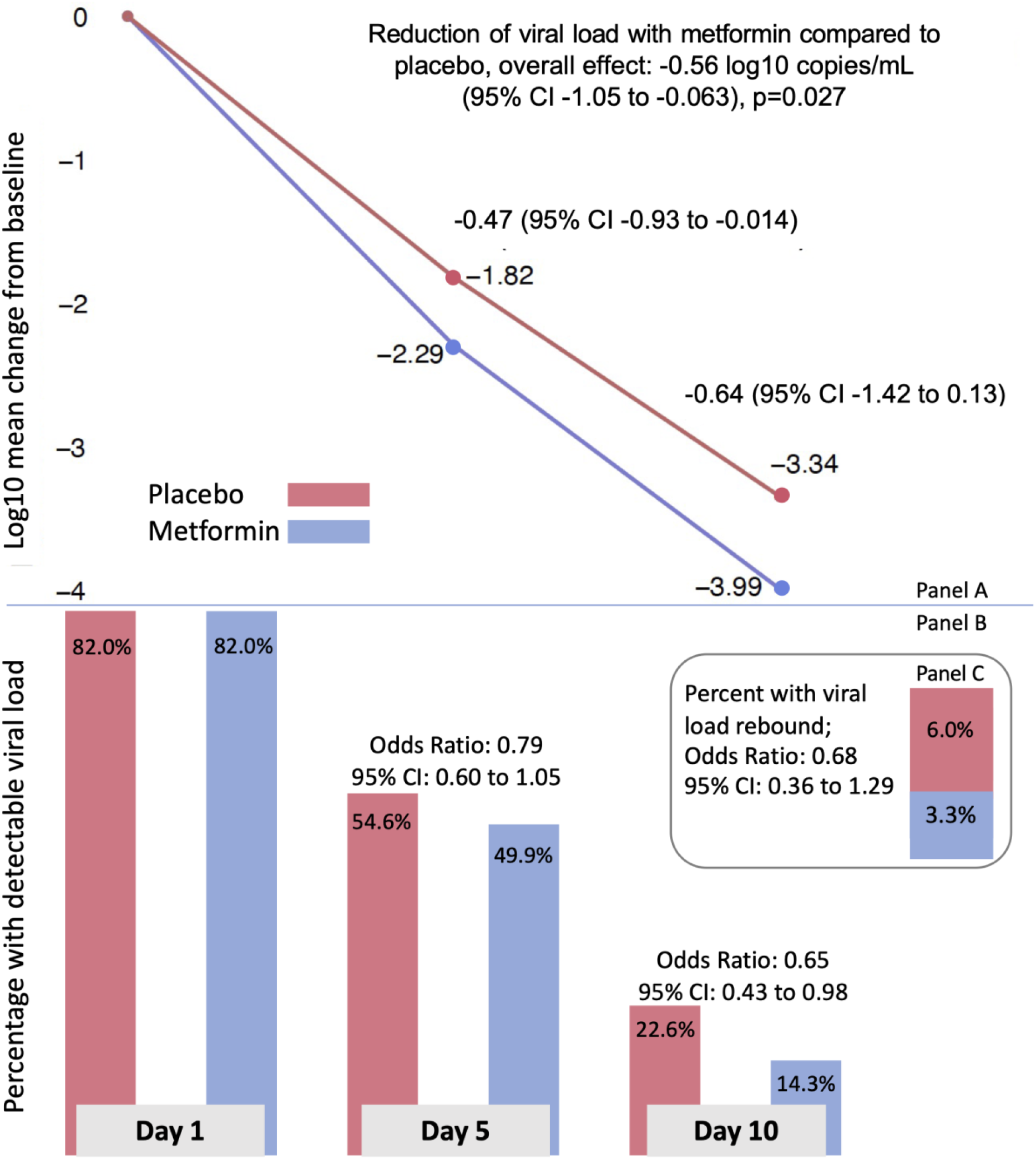
Effect of metformin versus placebo on viral load over time, detectable viral load, and rebound viral load. Panel A: adjusted mean change in log10 copies per ml (viral load) from baseline (Day 1) to Day 5 and Day 10 for metformin (blue) and placebo (red). Mean change estimates are based on the adjusted, multiply imputed Tobit analysis (the primary analytic approach) corresponding to the overall metformin analysis presented in Figure 2. Panel B: adjusted percent of viral load samples that were detectable at Day 1, 5, and 10. The percent viral load detected estimates were based on the adjusted, multiply imputed logistic GEE analysis corresponding to the overall metformin analysis depicted in Figure 3. Odds ratios correspond to adjusted effects on the odds ratio scale. Panel C: stacked bar chart depicting the adjusted percent of participants whose day 10 viral load was greater than the day 5 viral load, and the odds ratio for having viral load rebound.

**Figure 2.**
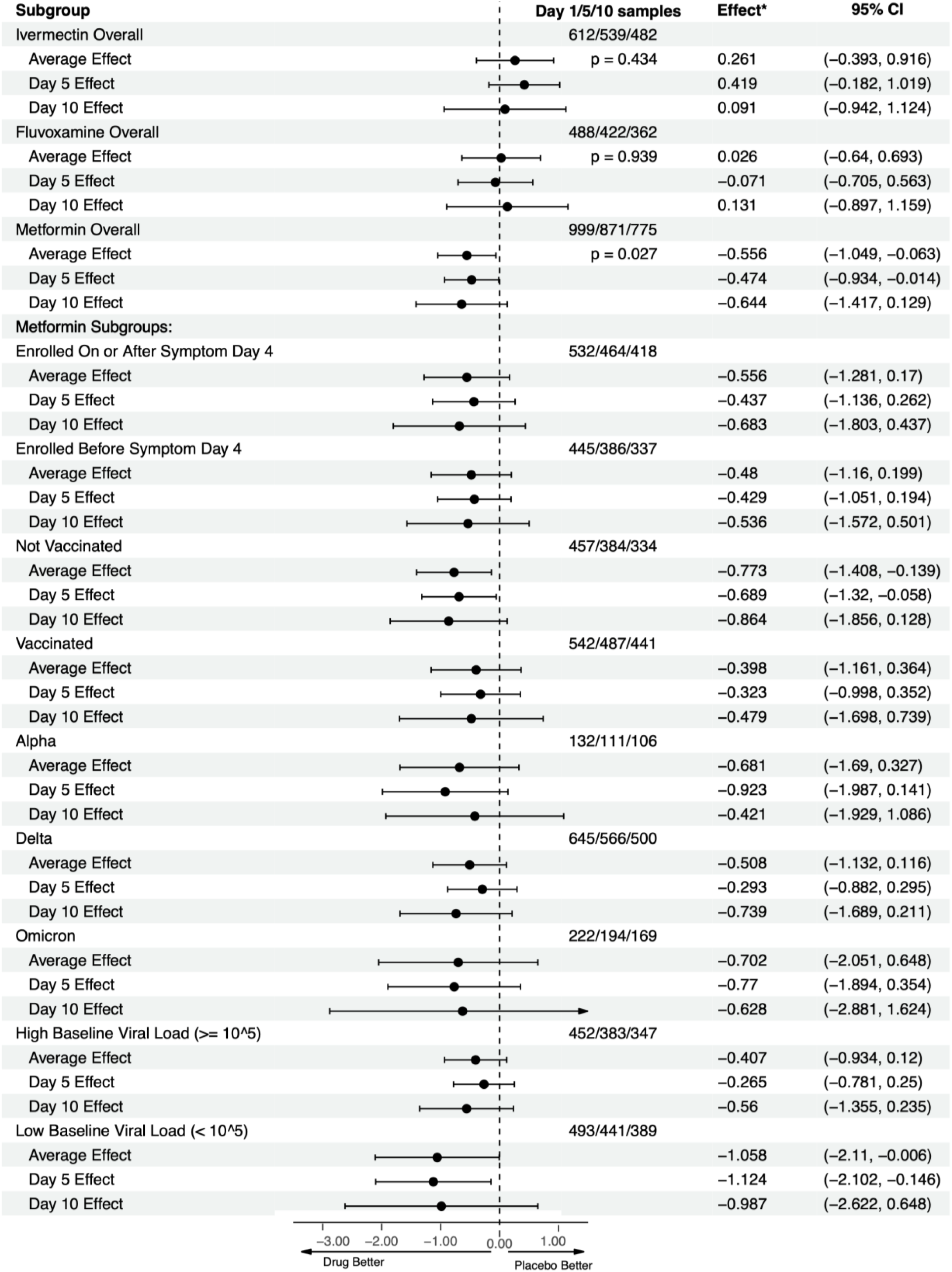
Overall results for metformin, ivermectin, and fluvoxamine on viral load, heterogeneity of treatment effect of metformin versus placebo. This is a forest plot that depicts the effect of active medication compared to control on log10 copies per ml (viral load), overall and at Days 5 and 10. “Viral Effect*” denotes the adjusted mean change in viral load in log10 copies per ml with 95% confidence intervals for the adjusted mean change. Analyses were conducted using the primary analytic approach, a multiply imputed Tobit model. The vertical dashed line indicates the value for a null effect. The top three rows show ivermectin, the next three rows show fluvoxamine, and the following three show metformin. Below these, the effect of metformin compared to placebo is shown by *a priori* subgroups of baseline characteristics.

When dropping the following adjustment covariates one at a time: baseline viral load, vaccination status, time since last vaccination, other study medications within the factorial trial, and the lab processing the nasal swabs in addition to dropping all adjustment covariates, the results were similar. The range in the estimated average effect was -0.51 log_10_ copies/mL (95% - 1.04 to 0.01, p=0.056) to -0.66 log_10_ copies/mL (95% CI -1.215 to -0.097, p=0.021) with the latter arising from the unadjusted model (**Table S8**).

Those in the metformin group were less likely to have a detectable viral load than placebo, (Odds Ratio (OR) 0.73; 95%CI, 0.58 to 0.96), **Figure 1**. This effect was higher at Day 10 (OR 0.65; 95%CI, 0.43 to 0.98), than Day 5 (OR 0.79; (95%CI, 0.60 to 1.05). Viral rebound was defined as having a higher viral load at Day 10 than Day 5. In the placebo group, 5.95% of participants had viral rebound, compared to 3.28% in the metformin group (OR 0.68; 95%CI, 0.36 to 1.29) for metformin compared to placebo, **Figure 1**.

Metformin’s effect on continuous viral load and conversion to undetectable viral load was consistent across *a priori* identified subgroups of baseline characteristics (**Figure 2, Figure 3**). Subgroups should be interpreted with caution because of low power, risk of making multiple comparisons without correction, and sparse data bias. ^13^ One subgroup warrants additional detail for interpretation: the antiviral effect on geometric log_10_ scale was greater among those with baseline viral loads <100,000 copies/mL (mean -1.17 log_10_ copies/mL reduction) than among those with >100,000 copies/mL (mean -0.49 log_10_ copies/mL reduction); although the reduction in absolute copies/mL would be greater among those with higher viral loads (**Figure 2, Figure 3)**.

**Figure 3.**
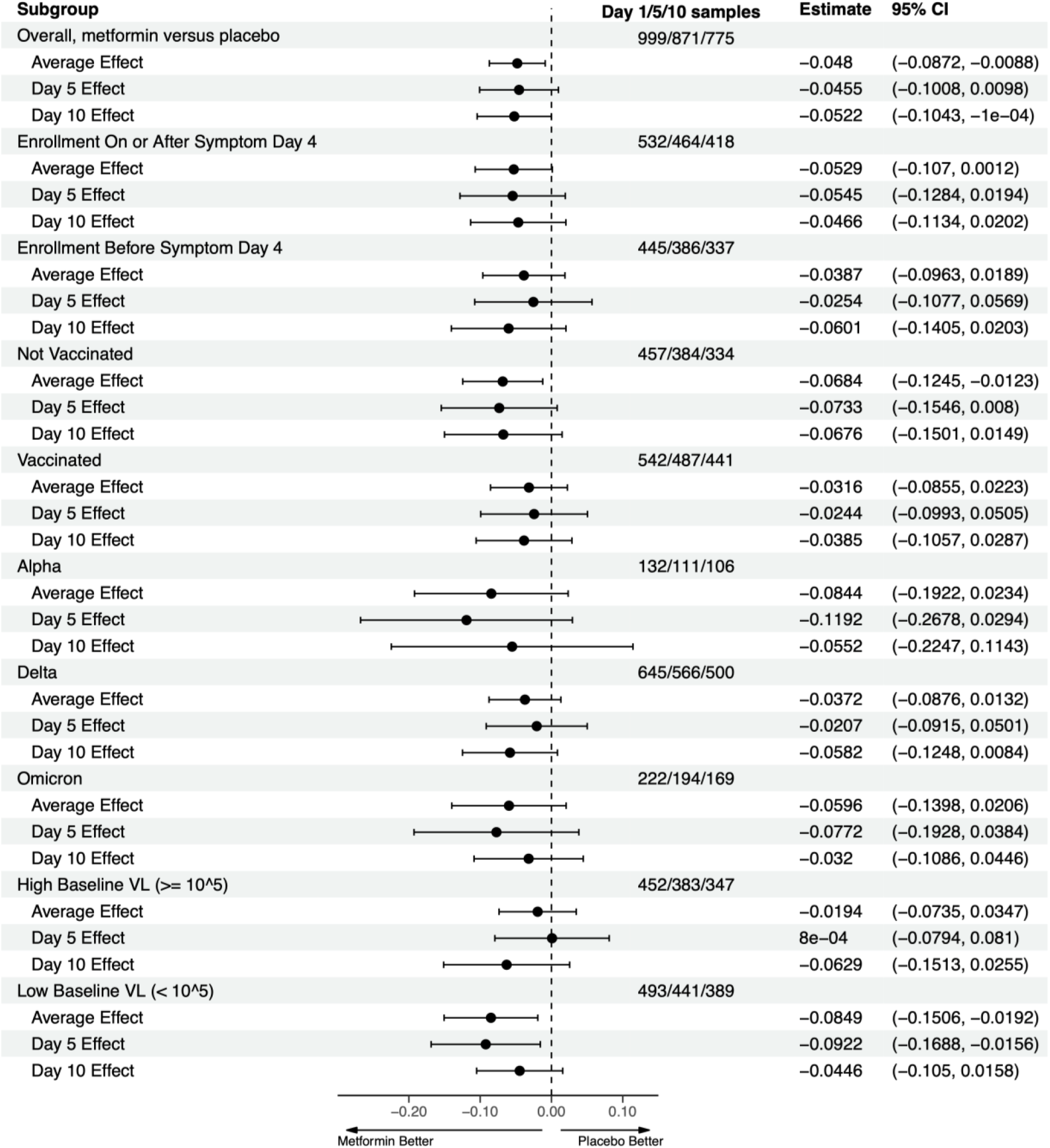
Overall results for metformin, ivermectin, and fluvoxamine on detectability of viral load; heterogeneity of treatment effect of metformin versus placebo. This is a forest plot that depicts the effect of active medication compared to control on the proportion of participants with a detectable viral load, overall and at Days 5 and 10. “Estimate” denotes the adjusted mean risk difference in the percent of samples with detected viral load with 95% confidence intervals for the adjusted risk difference. The vertical dashed line indicates the value for a null effect. The estimated risk differences are derived from the adjusted, multiply imputed logistic GEE analytic approach. The top three rows show ivermectin, the next three rows show fluvoxamine, and the following three show metformin. Below these, the effect of metformin compared to placebo is shown by *a priori* subgroups of baseline characteristics.

## Discussion

In the virologic endpoint of the COVID-OUT phase 3 randomized trial, metformin demonstrated a robust antiviral effect, reducing SARS-CoV-2 viral load over 10 days.^1^ The mean reduction was -0.56 log_10_ copies/mL greater than placebo. The antiviral response is consistent with the statistically significant and clinically relevant effects of metformin preventing clinical outcomes: severe COVID-19 (emergency room visit, hospitalization, or death) through Day 14; progression to hospitalization or death by Day 28; and the development of long COVID over 10 months. ^1,2^ The magnitude of effect on clinical outcomes was larger when metformin was started earlier in the course of infection, which is consistent with an antiviral effect.^1,2^

The antiviral effect in this phase 3 randomized trial is also consistent with emerging data from other trials. In a phase 2 randomized trial with 20 participants, the metformin group had better clinical outcomes, achieved an undetectable viral load 2.3 days faster than placebo (p=0.03); and a larger proportion of patients had an undetectable viral load at 3.3 days in the metformin group (p=0.04).^14^ A recent in vitro study showed that metformin decreased infectious SARS-CoV-2 titers and viral RNA in two cell lines, Caco2 and Calu3, at a clinically appropriate concentration.^15^

In the present trial, the magnitude of metformin’s antiviral effect was larger at Day 10 than Day 5 in the overall sample and across subgroups, which correlates with the dose titration to 1,500mg at Day 6. Previous work has shown metformin’s inhibition of mTOR complex 1 may depend on AMP-activated protein kinase (AMPK) at low doses but not high doses. ^6^ An AMPK-independent inhibition of mTOR may be more efficient. Additionally, metformin demonstrates a dose-dependent ability to inhibit IL-1, IL-6, and TNF-alpha in the presence of lipopolysaccharide (LPS), inflammatory products that correlate with COVID-19 severity. ^16,17^

The dose titration to 1,500mg over 6 days used in the COVID-OUT trial was faster than typical use. When used chronically for diabetes, prediabetes, or weight loss, metformin is slowly dose-titrated to 2,000mg daily over 4 to 8 weeks or longer. While metformin’s effect on diabetes control is not consistently dose-dependent, gastrointestinal side effects of metformin are dose-dependent. ^18^ Thus, despite what appears to be dose-dependent anti-viral and anti-inflammatory effects, a faster dose titration should likely only be considered in individuals with no side effects in the first days of metformin use.

In the context of other clinical investigations of SARS-CoV-2 antivirals, at Day 5 the antiviral effect over placebo was 0.47 log_10_ copies/mL for metformin, 0.30 log_10_ copies/mL for molnupiravir, and 0.80 log_10_ copies/mL for nirmatrelvir; and at Day 10 was 0.64 log_10_ copies/mL for metformin and 0.35 log_10_ copies/mL for nirmatrelvir in high risk participants. ^19,20^ We note that the trials enrolled different populations and at different times in the pandemic. When assessing for heterogeneity of effect, the metformin effect was consistent across subgroups. Metformin’s antiviral effect in vaccinated vs unvaccinated of -0.48 versus -0.86 log_10_ copies/mL at Day 10 mirrors nirmatrelvir, for which the effect in seropositive participants was smaller than the overall trial population, -0.13 versus -0.35 log_10_ copies/mL at Day 10. ^19^ Effective primed memory B and T cell anamnestic immunity prompting effective response by Day 5 in vaccinated persons may account for this trend in both trials. Subgroups should be interpreted with caution because of low power and multiple comparisons.^13^

Both nirmatrelvir and molnupiravir are pathogen-directed antiviral agents. There may be an important role for therapeutics that target host factors rather than viral factors, as targeting the host may be less likely to induce drug-resistant viral variants through mutation-selection.^21,22^ In addition to antiviral activity, metformin appears to have relevant anti-inflammatory actions. In mice without diabetes, metformin inhibited mitochondrial ATP and DNA synthesis to evade NLRP3 inflammasome activation.^9^ In macrophages of mice without diabetes infected with SARS-CoV-2, metformin inhibited inflammasome activation, IL-1 production, and IL-6 secretion, and also increased the IL-10 anti-inflammatory response to LPS, thereby attenuating LPS-induced lung injury.^23^ In a recent assay of human lung epithelial cell lines, metformin inhibited the cleavage of caspase-1 by NSP6, inhibiting the maturation and release of IL-1, a key factor mediating inflammatory responses.^8^ The idea of pleiotropic effects is being embraced in novel therapeutics being developed for both anti-viral and anti-inflammatory actions.^24^

Strengths of the study include the large sample size and detailed participant information collected, including the exact time and date of specimen collection. One limitation was the sampling time frame of only Day 1, 5, and 10 due to limited resources. By Day 10 post-randomization, 77% of participants in the placebo group had an undetectable viral load. As viral load is lower in vaccinated persons,^25^ this degree of undetectable viral loads differs from earlier clinical trials conducted in unvaccinated participants without known prior infection.^19,26^ Sampling earlier and more frequently, i.e. Days 1, 3, 6, and 9, may better characterize differences in viral shedding earlier in the infection and over time, dependent on the duration of therapy and timing of enrollment.

Future work could assess whether synergy exists between metformin and direct SARS-CoV-2 antivirals, as previous work showed metformin improved sustained virologic clearance of hepatitis C virus and improved outcomes in other respiratory infections.^27-29^ The biophysical modeling that motivated this trial predicts additive/cooperative effects in combination with protease inhibitors, and combination therapy might decrease resistance pressure caused by direct antivirals. Protease inhibitors interact with common medications, necessitating that those medications be held during a 5-day protease inhibitor treatment course. Metformin has few medication interactions, so treatment with metformin could continue beyond 5 days while home medications are restarted. Additionally, continuing metformin could reduce symptom rebound given its effects on T-cell immunity.^30,31^ Further data is needed to correlate whether decreased viral load and faster viral clearance decreases onward transmission of SARS-CoV-2. Additionally, this trial tested immediate release metformin, which reaches higher peak serum concentrations than extended-release, and it is unknown whether extended-release has the same anti-viral effects.^32^

## Conclusions

The demonstration that metformin has reproducible anti-SARS-CoV-2 activity in humans is an important development. The anti-viral effect in a phase 3 randomized trial provides the primary mechanism for the clinical benefits found in randomized trials of metformin to treat SARS-CoV-2. Metformin is a safe, inexpensive, and widely available medication that is over the counter in many countries. We have also confirmed that therapeutic targets can be rapidly identified using *in silico* modeling, which has wide-reaching implications beyond COVID-19. ^3^

## Data Availability

Data availability
The data for this manuscript, as well as the code used to generate the results, will be made freely available for individuals with a defined research question within approximately 1 month of it being published.

https://covidout.umn.edu

## Data availability

The data for this manuscript, as well as the code used to generate the results, will be made freely available for individuals with a defined research question within approximately 1 month of it being published.

## Author Contributions

All authors meet ICJME criteria for inclusion in the byline.

## Financial Disclosures

The fluvoxamine placebo tablets were donated by the Apotex pharmacy. The ivermectin placebo and active tablets were donated by the Edenbridge pharmacy. The trial was funded by the Parsemus Foundation, Rainwater Charitable Foundation, Fast Grants, and the UnitedHealth Group Foundation. The funders had no influence on the design or conduct of the trial and were not involved in data collection or analysis, writing of the manuscript, or decision to submit for publication. The authors assume responsibility for trial fidelity and the accuracy and completeness of the data and analyses.

Dr. Bramante was supported by grants (KL2TR002492 and UL1TR002494) from the National Center for Advancing Translational Sciences (NCATS) of the National Institutes of Health (NIH) and by a grant (K23 DK124654–01-A1) from the National Institute of Diabetes and Digestive and Kidney Diseases of the NIH. Dr. Buse was supported by a grant (UL1TR002489) from NCATS. Dr. Nicklas was supported by a grant (K23HL133604) from the National Heart, Lung, and Blood Institute of the NIH. Dr. Odde was supported by the Institute for Engineering in Medicine, UMN Office of Academic and Clinical Affairs COVID-19 Rapid Response Grant, the Earl E. Bakken Professorship for Engineering in Medicine, and by grants (U54 CA210190 and P01 CA254849) from the National Cancer Institute of the NIH. Dr. Murray was supported in part by the Medtronic Faculty Fellowship. Dr. Liebovitz receives funding from NIH RECOVER (OT2HL161847). Dr. Siegel was supported by NIH grants (18X107CF6 and 18X107CF5) through a contract with Leidos Biomedical and by grants from National Heart, Lung, and Blood Institute of the NIH (T32HL129956), and the NIH (R01LM012982 and R21LM012744). Dr. Puskarich receives grants from Bill and Melinda Gates Foundation (INV-017069), Minnesota Partnership for Biotechnology and Medical Genomics (00086722), National Heart, Lung, and Blood Institute of the NIH (OT2HL156812).

JBB reports contracted fees and travel support for contracted activities for consulting work paid to the University of North Carolina by Novo Nordisk; grant support by Dexcom, NovaTarg, Novo Nordisk, Sanofi, Tolerion and vTv Therapeutics; personal compensation for consultation from Alkahest, Altimmune, Anji, AstraZeneca, Bayer, Biomea Fusion Inc, Boehringer-Ingelheim, CeQur, Cirius Therapeutics Inc, Corcept Therapeutics, Eli Lilly, Fortress Biotech, GentiBio, Glycadia, Glyscend, Janssen, MannKind, Mellitus Health, Moderna, Pendulum Therapeutics, Praetego, Sanofi, Stability Health, Terns Inc, Valo and Zealand Pharma; and stock/options in Glyscend, Mellitus Health, Pendulum Therapeutics, PhaseBio, Praetego, and Stability Health. Dr. Puskarich receives consulting fees from Opticyte and Cytovale. Dr. Karger has served as an external consultant for Roche Diagnostics; received speaker honoraria from Siemens Healthcare Diagnostics, the American Kidney Fund, the National Kidney Foundation, and the American Society of Nephrology; and research support unrelated to this manuscript from Siemens Healthcare Diagnostics, Kyowa Kirin Pharmaceutical Development, the Juvenile Diabetes Research Foundation, and the NIH.

| **Quick-Reference BRISQ Checklist: Tier 1 Items.** |  |
| --- | --- |
| **Biospecimen type** | Nasal swab samples of Nasal mucous |
| **Anatomical site** | Anterior nares, mid-turbinate, bilaterally |
| **Clinical characteristics of patients** | Adults age 30-85 with a body mass index in the overweight or obesity category. |
| **Vital status of patients** | Alive at the time of collection |
| **Clinical diagnosis of patients** | Confirmed SARS-CoV-2 infection within less than 3 days |
| **Disease Status of patients** | Within less than 7 days of symptoms. |
| **Pathology diagnosis** | N/A |
| **Collection mechanism** | Participants self-collected with standardized instructions |
| **Type of stabilization** | Foam insulated shipping container with rigid cardboard and instant ice pack. |
| **Constitution of preservative** | Smart Transport Medium |
| **Storage temperature** | Approximately -2 to 10 degrees Celsius before receipt by the lab; then 4 degrees Celsius upon receipt at the lab |
| **Shipping temperature** | Approximately -2 to 20 degrees Celsius. |
| **Storage duration** | Approximately 1 to 4 days before receipt by the lab; then 1 to 3 days after receipt by the lab. |
| **Composition assessment and selection** | N/A |

Reference: https://acsjournals.onlinelibrary.wiley.com/doi/full/10.1002/cncy.20147

**CONSORT 2010 checklist of information to include when reporting a randomised trial\***

| Section/Topic | Item No | Checklist item | Reported on page No |
| --- | --- | --- | --- |
| **Title and abstract** |  |  |  |
|  | 1a | Identification as a randomised trial in the title | 1 |
|  | 1b | Structured summary of trial design, methods, results, and conclusions (for specific guidance see CONSORT for abstracts) | 1, 2 |
| **Introduction** |  |  |  |
| Background and objectives | 2a | Scientific background and explanation of rationale | 2 |
|  | 2b | Specific objectives or hypotheses | 2 |
| **Methods** |  |  |  |
| Trial design | 3a | Description of trial design (such as parallel, factorial) including allocation ratio | 1,2, Table S1,S9 |
|  | 3b | Important changes to methods after trial commencement (such as eligibility criteria), with reasons | Table S2 |
| Participants | 4a | Eligibility criteria for participants | 6 |
|  | 4b | Settings and locations where the data were collected | 6 |
| Interventions | 5 | The interventions for each group with sufficient details to allow replication, including how and when they were actually administered | 6 |
| Outcomes | 6a | Completely defined pre-specified primary and secondary outcome measures, including how and when they were assessed | 6, 7 |
|  | 6b | Any changes to trial outcomes after the trial commenced, with reasons | Table S2 |
| Sample size | 7a | How sample size was determined | Table S9 |
|  | 7b | When applicable, explanation of any interim analyses and stopping guidelines | n/a |
| Randomisation: |  |  |  |
| Sequence generation | 8a | Method used to generate the random allocation sequence | Table S9 |
|  | 8b | Type of randomisation; details of any restriction (such as blocking and block size) | Table S9 |
| Allocation concealment mechanism | 9 | Mechanism used to implement the random allocation sequence (such as sequentially numbered containers), describing any steps taken to conceal the sequence until interventions were assigned | Table S9 |
| Implementation | 10 | Who generated the random allocation sequence, who enrolled participants, and who assigned participants to interventions | Table S9 |
| Blinding | 11a | If done, who was blinded after assignment to interventions (for example, participants, care providers, those assessing outcomes) and how | 6 |
|  | 11b | If relevant, description of the similarity of interventions | 6 |
| Statistical methods | 12a | Statistical methods used to compare groups for primary and secondary outcomes | 6,7 |
|  | 12b | Methods for additional analyses, such as subgroup analyses and adjusted analyses | 6,7, Table S9 |
| **Results** |  |  |  |
| Participant flow (a diagram is strongly recommended) | 13a | For each group, the numbers of participants who were randomly assigned, received intended treatment, and were analysed for the primary outcome | Figure S1 |
|  | 13b | For each group, losses and exclusions after randomisation, together with reasons | Figure S1 |
| Recruitment | 14a | Dates defining the periods of recruitment and follow-up | 6 |
|  | 14b | Why the trial ended or was stopped | 6 |
| Baseline data | 15 | A table showing baseline demographic and clinical characteristics for each group | Table 1 |
| Numbers analysed | 16 | For each group, number of participants (denominator) included in each analysis and whether the analysis was by original assigned groups | Table 1, Figure S1 |
| Outcomes and estimation | 17a | For each primary and secondary outcome, results for each group, and the estimated effect size and its precision (such as 95% confidence interval) | Figures 1, 2, 3 |
|  | 17b | For binary outcomes, presentation of both absolute and relative effect sizes is recommended |  |
| Ancillary analyses | 18 | Results of any other analyses performed, including subgroup analyses and adjusted analyses, distinguishing pre-specified from exploratory | Supplement |
| Harms | 19 | All important harms or unintended effects in each group (for specific guidance see CONSORT for harms) | n/a |
| **Discussion** |  |  |  |
| Limitations | 20 | Trial limitations, addressing sources of potential bias, imprecision, and, if relevant, multiplicity of analyses | 5 |
| Generalisability | 21 | Generalisability (external validity, applicability) of the trial findings | 5 |
| Interpretation | 22 | Interpretation consistent with results, balancing benefits and harms, and considering other relevant evidence | 5 |
| **Other information** |  |  |  |
| Registration | 23 | Registration number and name of trial registry | 6 |
| Protocol | 24 | Where the full trial protocol can be accessed, if available | clinicaltrials.gov |
| Funding | 25 | Sources of funding and other support (such as supply of drugs), role of funders | 8 |

\*We strongly recommend reading this statement in conjunction with the CONSORT 2010 Explanation and Elaboration for important clarifications on all the items. If relevant, we also recommend reading CONSORT extensions for cluster randomised trials, non-inferiority and equivalence trials, non-pharmacological treatments, herbal interventions, and pragmatic trials. Additional extensions are forthcoming; for those and for up to date references relevant to this checklist, see www.consort-statement.org.

## Notes

### Competing Interest Statement

The authors have declared no competing interest.

### Clinical Trial

ClinicalTrials.gov: NCT04510194

### Author Declarations

The Institutional Review Board of Advarra gave ethical approval for this work (MET29324).

