## Supplemental Appendix for "Metformin reduces SARS-CoV-2 in a Phase 3 Randomized Placebo Controlled Clinical Trial"

**Supplementary Material, Table of Contents.**


| Item | Description | Page |
|---|---|---|
| Table S1 | COVID-OUT Study Team | 2-3 |
| Table S2 | Overview of the factorial design of the trial | 4 |
| Figure S1 | Consort of participants in the Viral load analysis. | 5 |
| **Demographics of participants submitting any nasal swab versus no nasal swab** | | |
| Table S3 | Table 1, those submitting any swabs versus no swabs, metformin | 6 |
| Table S4 | Table 1, those submitting any swabs versus no swabs, ivermectin | 7 |
| Table S5 | Table 1, those submitting any swabs versus no swabs, fluvoxamine | 8 |
| **Demographics of participants submitting nasal swabs on Day 1, Day 5, and Day 10.** | | |
| Table S6 | Table 1 by Day 1, Day 5, and Day 10, metformin. | 9 |
| **Additional results** | | |
| Table S7 | Percent undetectable, all medications. | 10 |
| Table S8 | Sequentially dropping adjustment covariates | 10 |
| **Complete Case Analyses** | | |
| Figure S2 | Change in viral load from baseline to follow-up, all three medications. | 11 |
| Figure S3 | Percent Detectable and Rebound, metformin versus placebo | 12 |
| Figure S4 | Viral load over time, metformin versus placebo | 13 |
| **Additional study details** | | |
| Table S9 | Additional detail on analytic approach | 14 |
| Table S10 | Additional detail on laboratory procedures | 14 |
| **Sensitivity Analyses** | | |
| Figure S5 | Results for each lab separately | 15 |
| Figure S6 | Viral load over time, manual imputation | 16 |
| Figure S7 | Viral load over time, imputed from level of quantification | 17 |
| Table S11 | Excluding participants who obtained first swab after taking study drug | 18 |

**Table S1. COVID-OUT Study Team**

| Name | Institute | Location |
|---|---|---|
| Blake Anderson | Emory University | Atlanta, GA |
| Riannon C Atwater | University of Colorado | Aurora, CO |
| Nandini Avula | University of Minnesota | Minneapolis, MN |
| Kenny B Beckman | University of Minnesota | Minneapolis, MN |
| Hrishikesh K Belani | Olive View - UCLA | Sylmar, CA |
| David R Boulware | University of Minnesota | Minneapolis, MN |
| Carolyn T Bramante | University of Minnesota | Minneapolis, MN |
| Jannis Brea | Northwestern University | Chicago, IL |
| Courtney A Broedlow | University of Minnesota | Minneapolis, MN |
| John B Buse | University of North Carolina | Chapel Hill, NC |
| Paula Campora | University of Minnesota | Minneapolis, MN |
| Anup Challa | Vanderbilt University | Nashville, TN |
| Jill Charles | University of Minnesota | Minneapolis, MN |
| Grace Christensen | University of Minnesota | Minneapolis, MN |
| Theresa Christiansen | M Health Fairview | Minneapolis, MN |
| Ken Cohen | Optum | Minnetonka, MN |
| Bo Connelly | University of Minnesota | Minneapolis, MN |
| Srijani Datta | University of Minnesota | Minneapolis, MN |
| Nikita Deng | University of Colorado | Aurora, CO |
| Alex T Dunn | Hennepin Healthcare | Minneapolis, MN |
| Spencer M Erickson | University of Minnesota | Minneapolis, MN |
| Faith M Fairbairn | University of Minnesota | Minneapolis, MN |
| Sarah L Fenno | University of Minnesota | Minneapolis, MN |
| Daniel J Fraser | University of Minnesota | Minneapolis, MN |
| Regina D Fricton | Feinberg School of Medicine, Northwestern | Chicago, IL |
| Gwen Griffiths | University of Minnesota | Minneapolis, MN |
| Aubrey A Hagen | University of Minnesota | Minneapolis, MN |
| Katrina M Hartman | University of Minnesota | Minneapolis, MN |
| Audrey F Hendrickson | Hennepin Healthcare | Minneapolis, MN |
| Jared D Huling | University of Minnesota | Minneapolis, MN |
| Nicholas E Ingraham | University of Minnesota | Minneapolis, MN |
| Arthur C Jeng | Olive View - UCLA | Sylmar, CA |
| Darrell M Johnson | University of Minnesota | Minneapolis, MN |
| Amy B Karger | University of Minnesota | Minneapolis, MN |
| Nichole R Klatt | University of Minnesota | Minneapolis, MN |
| Erik A Kuehl | M Health Fairview | Minneapolis, MN |
| Derek D LaBar | M Health Fairview | Minneapolis, MN |
| Samuel Lee | Feinberg School of Medicine, Northwestern | Chicago, IL |
| David M Liebovitz | Feinberg School of Medicine, Northwestern | Chicago, IL |

| | | |
|---|---|---|
| Sarah Lindberg | University of Minnesota | Minneapolis, MN |
| Darlette G Luke | M Health Fairview | Minneapolis, MN |
| Rosario Machicado | Olive View - UCLA | Sylmar, CA |
| Zeinab Mohamud | University of Minnesota | Minneapolis, MN |
| Thomas A Murray | University of Minnesota | Minneapolis, MN |
| Rumbidzai Ngonyama | University of Minnesota | Minneapolis, MN |
| Jacinda M Nicklas | University of Colorado | Aurora, CO |
| David J Odde | University of Minnesota | Minneapolis, MN |
| Elliott Parrens | M Health Fairview | Minneapolis, MN |
| Daniela Parra | University of Minnesota | Minneapolis, MN |
| Barkha Patel | University of Minnesota | Minneapolis, MN |
| Jennifer L Proper | University of Minnesota | Minneapolis, MN |
| Matthew F Pullen | University of Minnesota | Minneapolis, MN |
| Michael A Puskarich | Hennepin Healthcare | Minneapolis, MN |
| Via Rao | University of Minnesota | Minneapolis, MN |
| Neha V Reddy | University of Minnesota | Minneapolis, MN |
| Naveen Reddy | Northwestern University | Chicago, IL |
| Katelyn J Rypka | University of Minnesota | Minneapolis, MN |
| Hanna G Saveraid | University of Minnesota | Minneapolis, MN |
| Paula Seloadji | Olive View - UCLA | Sylmar, CA |
| Arman Shahriar | University of Minnesota | Minneapolis, MN |
| Nancy Sherwood | University of Minnesota | Minneapolis, MN |
| Jamie L Siegart | University of Colorado | Aurora, CO |
| Lianne K Siegel | University of Minnesota | Minneapolis, MN |
| Lucas Simmons | University of Minnesota | Minneapolis, MN |
| Isabella Sinelli | University of Colorado | Aurora, CO |
| Palak Singh | University of Minnesota | Minneapolis, MN |
| Andrew Snyder | M Health Fairview | Minneapolis, MN |
| Maxwell T Stauffer | St. Olaf College | Northfield, MN |
| Jennifer Thompson | Vanderbilt University | Nashville, TN |
| Christopher J Tignanelli | University of Minnesota | Minneapolis, MN |
| Tannon L Tople | University of Minnesota | Minneapolis, MN |
| Walker J Tordsen | Hennepin Healthcare | Minneapolis, MN |
| Ray HB Watson | University of Minnesota | Minneapolis, MN |
| Beiqing Wu | University of Minnesota | Minneapolis, MN |
| Adnin Zaman | University of Colorado | Aurora, CO |
| Madeline R Zolik | M Health Fairview | Minneapolis, MN |
| Lena Zinkl | M Health Fairview | Minneapolis, MN |

Back to Top

| **Table S2. Overview of factorial design groups.** | | |
|---|---|---|
| | **Metformin** | **Metformin Placebo** |
| Ivermectin | **1:** Metformin + Ivermectin | **4:** Metformin Placebo + Ivermectin |
| Fluvoxamine | **2:** Metformin + Fluvoxamine | **5:** Metformin Placebo + Fluvoxamine |
| Placebo (Ivermectin or Fluvoxamine) | **3:** Metformin + Placebo | **6:** Metformin Placebo + Placebo |

**Metformin trial:** groups 1 + 2 + 3 **vs** groups 4 + 5 + 6

**Fluvoxamine trial:** groups 2 + 5 **vs** groups 3 + 6

**Ivermectin trial:** groups 1 + 4 **vs** groups 3 + 6

Adjustment for multi-comparisons is not indicated for having three medications assessed in a parallel-group factorial design trial.[a] In this 2x3 factorial design, groups 1 and 2 had two active medications. Therefore, each participant received two types of study pills to maintain the blind.

Every participant in the trial received a pill that looked like metformin – either active metformin or exact-matching metformin placebo. The study started with just metformin versus placebo and then expanded to the factorial randomization of parallel arms to include fluvoxamine and ivermectin because of the importance of having phase 3 clinical trial results for those medications and the ability to study all 3 in an efficient way while maintaining the blind.[b]

The second pill was either ivermectin or exact-matching ivermectin placebo; or fluvoxamine or exact-matching fluvoxamine placebo. A small subset of the control group for fluvoxamine received ivermectin placebo, and a small subset of the control group for ivermectin received the fluvoxamine placebo, because of shipping and supply chain issues. For that reason, the control groups for fluvoxamine and ivermectin are referred to as control or blinded control rather than placebo like metformin.

Pills were dispensed in pre-filled pill boxes to assure the right number of each pill was taken.[b]

[a] Parker RA, Weir CJ. Non-adjustment for multiple testing in multi-arm trials of distinct treatments: Rationale and justification. *Clinical Trials*. 2020;17(5):562-566. doi:10.1177/1740774520941419

[b] Bramante CT, Huling JD, Tignanelli CJ, et al. Randomized Trial of Metformin, Ivermectin, and Fluvoxamine for Covid-19. *The New England journal of medicine*. Aug 18 2022;387(7):599-610. doi:10.1056/NEJMoa2201662

Back to Top

**Figure S1. This is a CONSORT diagram of participants included in the viral load analysis.**

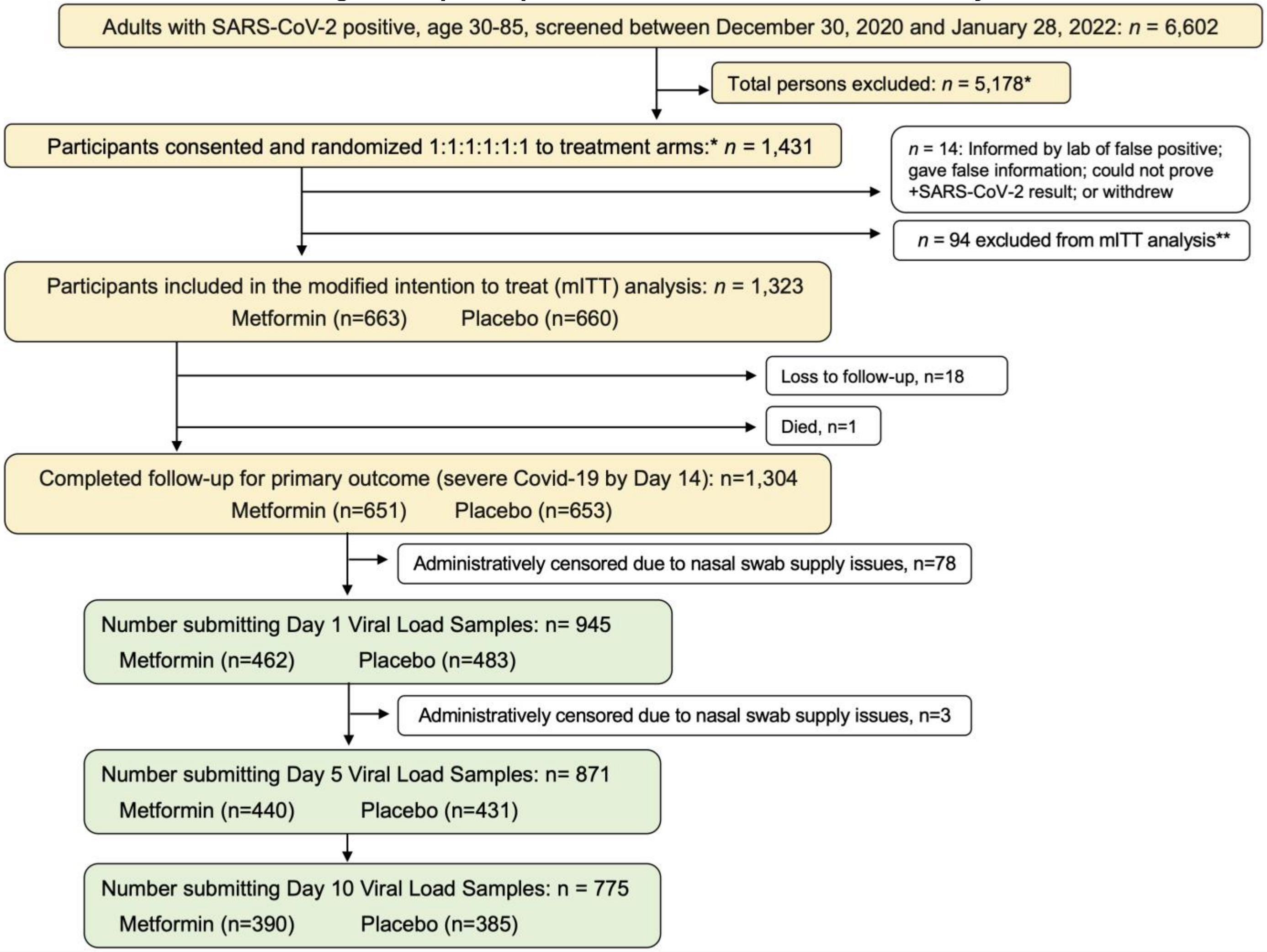


The primary analytic sample for the trial was decided *a priori* to be a modified intention to treat (mITT) sample because of the remote nature of the study. *The eligibility exclusions have been published. **Participants were excluded from the mITT sample if they did take any study pills: did not receive the medication (n=9); were hospitalized at the time the medication arrived (n=8); or by the time they received the medication they had changed their mind about willingness to participate in the study protocol by taking study pills (n=77). Only 3 participants in the full intention to treat sample but not in the mITT sample submitted nasal swabs. All results are presented in the mITT group, the *a priori* primary sample. Back to Top

**Table S3**. **Demographic characteristics of persons who submitted any nasal swab versus no nasal swab.**

| | | Submitted Any Nasal Swab | | | Submitted No Nasal Swab | | |
|---|---|---|---|---|---|---|---|
| **Variable** | | Overall N = 999[1] | Control N = 495[1] | Metformin N = 504[1] | Overall N = 324[1] | Control N = 165[1] | Metformin N = 159[1] |
| **Age** | | 46 (38, 55) | 45 (38, 54) | 46 (38, 55) | 44 (36, 54) | 45 (36, 55) | 44 (37, 53) |
| **Biologic Sex,** Female**,** %(N) | | 56% (559) | 57% (282) | 55% (277) | 56% (182) | 61% (100) | 52% (82) |
| **Race** | Native American | 2.2% (22) | 2.6% (13) | 1.8% (9) | 1.5% (5) | 2.4% (4) | 0.6% (1) |
| | Asian | 3.6% (36) | 3.8% (19) | 3.4% (17) | 4.6% (15) | 4.2% (7) | 5.0% (8) |
| | Native. Haw. or Pac. Isl. | 0.7% (7) | 0.4% (2) | 1.0% (5) | 0.6% (2) | 1.2% (2) | 0% (0) |
| | Black or African American | 6.2% (62) | 6.1% (30) | 6.3% (32) | 12% (38) | 9.1% (15) | 14% (23) |
| | White | 85% (849) | 85% (420) | 85% (429) | 75% (242) | 76% (126) | 73% (116) |
| | Other/Declined | 4.3% (43) | 3.8% (19) | 4.8% (24) | 8.3% (27) | 7.3% (12) | 9.4% (15) |
| | Race Missing | 0.7% (7) | 0.6% (3) | 0.8% (4) | 0.9% (3) | 1.8% (3) | 0% (0) |
| **Hispanic** | | 12% (118) | 13% (63) | 11% (55) | 13% (42) | 13% (21) | 13% (21) |
| Unknown | | 0.6% (6) | 0.8% (4) | 0.4% (2) | 1.9% (6) | 3.0% (5) | 0.6% (1) |
| **Time since last vaccine dose (days)** | | 194.0 (132.2, 240.0) | 195.0 (132.5, 234.5) | 192.0 (132.5, 245.5) | 172.0 (116.0, 230.2) | 160.5 (116.5, 231.2) | 178.0 (116.0, 226.0) |
| **Vaccination status at baseline** | No Vaccine | 46% (457) | 48% (240) | 43% (217) | 62% (200) | 61% (101) | 62% (99) |
| | Primary Series Only | 50% (495) | 47% (232) | 52% (263) | 35% (115) | 36% (60) | 35% (55) |
| | Booster | 4.7% (47) | 4.6% (23) | 4.8% (24) | 2.8% (9) | 2.4% (4) | 3.1% (5) |
| **Medical History** | BMI | 30.0 (27.1, 34.3) | 30.0 (26.9, 34.7) | 29.8 (27.2, 34.0) | 29.5 (26.6, 33.7) | 29.8 (26.5, 33.9) | 29.4 (26.8, 33.4) |
| | BMI >= 30kg/m2 | 50% (496) | 51% (250) | 49% (246) | 46% (150) | 48% (80) | 44% (70) |
| | Cardiovascular disease | 28% (282) | 28% (140) | 28% (142) | 22% (71) | 21% (35) | 23% (36) |
| | Diabetes | 2.0% (20) | 2.6% (13) | 1.4% (7) | 1.9% (6) | 1.8% (3) | 1.9% (3) |
| **Symptom duration on initiation (days)** | | 4.7 (1.9) | 4.7 (1.8) | 4.7 (1.9) | 4.9 (2.1) | 4.8 (2.2) | 4.9 (2.0) |
| **Symptom duration <= 4 days** | | 46% (453) | 48% (230) | 45% (223) | 48% (151) | 48% (76) | 47% (75) |
| **Variant Period** | Alpha | 13% (132) | 13% (65) | 13% (67) | 8.3% (27) | 9.1% (15) | 7.5% (12) |
| | Delta | 65% (645) | 65% (320) | 64% (325) | 70% (226) | 67% (111) | 72% (115) |
| | Omicron | 22% (222) | 22% (110) | 22% (112) | 22% (71) | 24% (39) | 20% (32) |
| **Insurance Type** | Private | 65% (652) | 65% (324) | 65% (328) | 53% (171) | 54% (89) | 52% (82) |
| | Medicare | 7.5% (75) | 6.9% (34) | 8.1% (41) | 7.7% (25) | 8.5% (14) | 6.9% (11) |
| | Medicaid | 14% (136) | 14% (69) | 13% (67) | 20% (64) | 24% (39) | 16% (25) |
| | No insurance | 12% (123) | 12% (60) | 12% (63) | 17% (55) | 13% (21) | 21% (34) |
| | Unknown | 1.3% (13) | 1.6% (8) | 1.0% (5) | 2.8% (9) | 1.2% (2) | 4.4% (7) |

[1]Median (IQR); % (n); Mean (SD) Abbreviation: BMI=body mass index

Back to Top

**Table S4. Demographic characteristics of persons who submitted any nasal swab versus no nasal swab, ivermectin.**

| | | Submitted Any Nasal Swab | | | Submitted No Nasal Swab | | |
|---|---|---|---|---|---|---|---|
| **Variable** | | Overall, N = 612[1] | Control, N = 293[1] | Ivermectin, N = 319[1] | Overall, N = 196[1] | Control, N = 105[1] | Ivermectin, N = 91[1] |
| **Age** | | 46 (38, 55) | 45 (37, 56) | 47 (40, 55) | 45 (37, 55) | 45 (37, 56) | 45 (37, 54) |
| **Biologic Sex,** Female**,** %(N) | | 55% (335) | 57% (167) | 53% (168) | 55% (107) | 56% (59) | 53% (48) |
| **Race** | Native American | 2.0% (12) | 2.4% (7) | 1.6% (5) | 2.0% (4) | 1.9% (2) | 2.2% (2) |
| | Asian | 4.2% (26) | 4.4% (13) | 4.1% (13) | 5.6% (11) | 4.8% (5) | 6.6% (6) |
| | Native. Haw. or Pac. Isl. | 0.5% (3) | 0.7% (2) | 0.3% (1) | 1.0% (2) | 1.0% (1) | 1.1% (1) |
| | Black or African American | 5.9% (36) | 6.1% (18) | 5.6% (18) | 12% (23) | 10% (11) | 13% (12) |
| | White | 85% (522) | 84% (245) | 87% (277) | 71% (140) | 73% (77) | 69% (63) |
| | Other/Declined | 4.6% (28) | 5.5% (16) | 3.8% (12) | 9.7% (19) | 10% (11) | 8.8% (8) |
| | Race Missing | 0.7% (4) | 0.3% (1) | 0.9% (3) | 1.0% (2) | 1.0% (1) | 1.1% (1) |
| **Hispanic** | | 12% (72) | 15% (44) | 8.8% (28) | 13% (26) | 12% (13) | 14% (13) |
| Unknown | | 0.7% (4) | 1.4% (4) | 0% (0) | 2.6% (5) | 2.9% (3) | 2.2% (2) |
| **Time since last vaccine dose (days)** | | 195.5 (147.2, 248.0) | 191.5 (145.5, 248.0) | 197.5 (151.0, 248.8) | 177.0 (124.5, 231.5) | 155.0 (117.0, 232.0) | 208.0 (146.0, 231.0) |
| **Vaccination status at baseline** | No Vaccine | 42% (254) | 39% (113) | 44% (141) | 60% (117) | 61% (64) | 58% (53) |
| | Primary Series Only | 53% (327) | 56% (163) | 51% (164) | 38% (74) | 36% (38) | 40% (36) |
| | Booster | 5.1% (31) | 5.8% (17) | 4.4% (14) | 2.6% (5) | 2.9% (3) | 2.2% (2) |
| **Medical History** | BMI | 29.7 (27.1, 33.7) | 29.6 (26.9, 33.7) | 29.8 (27.2, 33.7) | 29.3 (26.5, 33.9) | 29.6 (26.6, 34.3) | 28.9 (26.5, 32.3) |
| | BMI >= 30kg/m2 | 49% (297) | 48% (140) | 49% (157) | 43% (85) | 46% (48) | 41% (37) |
| | Cardiovascular disease | 24% (145) | 22% (65) | 25% (80) | 20% (40) | 23% (24) | 18% (16) |
| | Diabetes | 1.8% (11) | 1.4% (4) | 2.2% (7) | 1.5% (3) | 1.9% (2) | 1.1% (1) |
| **Symptom duration on initiation (days)** | | 4.7 (1.8) | 4.8 (1.8) | 4.6 (1.8) | 4.7 (2.0) | 4.9 (1.9) | 4.6 (2.1) |
| **Symptom duration <= 4 days** | | 46% (278) | 46% (132) | 46% (146) | 48% (94) | 39% (41) | 59% (53) |
| **Variant Period** | Alpha | 2.8% (17) | 2.4% (7) | 3.1% (10) | 2.6% (5) | 3.8% (4) | 1.1% (1) |
| | Delta | 68% (418) | 69% (202) | 68% (216) | 69% (135) | 70% (73) | 68% (62) |
| | Omicron | 29% (177) | 29% (84) | 29% (93) | 29% (56) | 27% (28) | 31% (28) |
| **Insurance Type** | Private | 64% (392) | 62% (183) | 66% (209) | 48% (95) | 45% (47) | 53% (48) |
| | Medicare | 6.7% (41) | 7.8% (23) | 5.6% (18) | 8.7% (17) | 7.6% (8) | 9.9% (9) |
| | Medicaid | 15% (90) | 13% (39) | 16% (51) | 20% (40) | 20% (21) | 21% (19) |
| | No insurance | 13% (81) | 14% (42) | 12% (39) | 19% (38) | 24% (25) | 14% (13) |
| | Unknown | 1.3% (8) | 2.0% (6) | 0.6% (2) | 3.1% (6) | 3.8% (4) | 2.2% (2) |

[1]Median (IQR); % (n); Mean (SD) Abbreviation: BMI=body mass index

Back to Top

**Table S5. Demographic characteristics of persons who submitted any nasal swab versus no nasal swab, fluvoxamine.**

| | | Submitted Any Nasal Swab | | | Submitted No Nasal Swab | | |
|---|---|---|---|---|---|---|---|
| | | Overall, n = 488[1] | Control, n = 240[1] | Fluvoxamine, n =248[1] | Overall, n = 173[1] | Control, n = 87[1] | Fluvoxamine, n = 86[1] |
| **Age** | | 45 (37, 53) | 44 (37, 53) | 46 (39, 53) | 44 (37, 53) | 42 (37, 52) | 45 (37, 53) |
| **Biologic Sex,** Female**,** %(N) | | 55% (270) | 58% (140) | 52% (130) | 51% (88) | 55% (48) | 47% (40) |
| **Race** | Native American | 2.9% (14) | 2.9% (7) | 2.8% (7) | 1.7% (3) | 2.3% (2) | 1.2% (1) |
| | Asian | 3.3% (16) | 4.2% (10) | 2.4% (6) | 2.9% (5) | 2.3% (2) | 3.5% (3) |
| | Native. Haw. or Pac. Isl. | 0.8% (4) | 0.8% (2) | 0.8% (2) | 0.6% (1) | 1.1% (1) | 0% (0) |
| | Black or African American | 6.6% (32) | 5.4% (13) | 7.7% (19) | 11% (19) | 11% (10) | 10% (9) |
| | White | 83% (406) | 85% (203) | 82% (203) | 77% (133) | 74% (64) | 80% (69) |
| | Other/Declined | 4.7% (23) | 5.0% (12) | 4.4% (11) | 8.7% (15) | 10% (9) | 7.0% (6) |
| | Race Missing | 0.8% (4) | 0.4% (1) | 1.2% (3) | 1.2% (2) | 1.1% (1) | 1.2% (1) |
| **Hispanic** | | 14% (67) | 15% (35) | 13% (32) | 12% (21) | 13% (11) | 12% (10) |
| Unknown | | 1.0% (5) | 1.7% (4) | 0.4% (1) | 2.3% (4) | 3.4% (3) | 1.2% (1) |
| **Time since last vaccine dose (days)** | | 191.0 (135.5, 230.5) | 180.0 (134.0, 224.5) | 198.5 (135.8, 234.2) | 154.5 (108.8, 217.5) | 147.0 (113.0, 204.0) | 162.0 (106.0, 220.0) |
| **Vaccination status at baseline** | No Vaccine | 39% (189) | 37% (89) | 40% (100) | 61% (105) | 62% (54) | 59% (51) |
| | Primary Series Only | 57% (277) | 57% (137) | 56% (140) | 36% (62) | 36% (31) | 36% (31) |
| | Booster | 4.5% (22) | 5.8% (14) | 3.2% (8) | 3.5% (6) | 2.3% (2) | 4.7% (4) |
| **Medical History** | BMI | 29.4 (27.0, 34.3) | 29.5 (27.2, 33.9) | 29.3 (26.9, 34.4) | 29.6 (27.0, 34.0) | 29.6 (26.7, 35.2) | 29.6 (27.2, 33.1) |
| | BMI >= 30kg/m2 | 47% (229) | 48% (116) | 46% (113) | 47% (82) | 47% (41) | 48% (41) |
| | Cardiovascular disease | 28% (135) | 22% (53) | 33% (82) | 24% (42) | 24% (21) | 24% (21) |
| | Diabetes | 1.0% (5) | 0.4% (1) | 1.6% (4) | 1.2% (2) | 2.3% (2) | 0% (0) |
| **Symptom duration on initiation (days)** | | 4.8 (1.9) | 4.7 (1.8) | 4.9 (2.1) | 5.0 (2.2) | 4.9 (2.0) | 5.0 (2.4) |
| **Symptom duration <= 4 days** | | 46% (218) | 48% (113) | 43% (105) | 43% (75) | 38% (33) | 49% (42) |
| **Variant Period** | Alpha | 3.5% (17) | 2.9% (7) | 4.0% (10) | 3.5% (6) | 4.6% (4) | 2.3% (2) |
| | Delta | 83% (404) | 84% (202) | 81% (202) | 86% (148) | 84% (73) | 87% (75) |
| | Omicron | 14% (67) | 13% (31) | 15% (36) | 11% (19) | 11% (10) | 10% (9) |
| **Insurance Type** | Private | 63% (309) | 64% (154) | 62% (155) | 54% (94) | 49% (43) | 59% (51) |
| | Medicare | 8.0% (39) | 6.7% (16) | 9.3% (23) | 5.2% (9) | 5.7% (5) | 4.7% (4) |
| | Medicaid | 12% (57) | 12% (28) | 12% (29) | 16% (27) | 16% (14) | 15% (13) |
| | No insurance | 15% (75) | 15% (37) | 15% (38) | 21% (37) | 24% (21) | 19% (16) |
| | Unknown | 1.6% (8) | 2.1% (5) | 1.2% (3) | 3.5% (6) | 4.6% (4) | 2.3% (2) |

[1]Median (IQR); % (n); Mean (SD). Abbreviation: BMI=body mass index

Back to Top

| Table S6. Demographic characteristics of those who submitted nasal samples at each time point, overall and by metformin. | | | | | | | | | | |
|---|---|---|---|---|---|---|---|---|---|---|
| | | Submitted Day 1 | | | Submitted Day 5 | | | Submitted Day 10 | | |
| | | Overall, N = 945[1] | Control, N = 462[1] | Metformin, N = 483[1] | Overall, N = 871[1] | Control, N = 440[1] | Metformin, N = 431[1] | Overall, N = 775[1] | Control, N = 390[1] | Metformin, N = 385[1] |
| **Age** | | 46 (38, 55) | 45 (37, 54) | 46 (38, 55) | 46 (38, 55) | 46 (38, 55) | 46 (38, 56) | 46 (38, 56) | 46 (38, 56) | 46 (38, 56) |
| **Biologic Sex,** Female, %(N) | | 56% (529) | 58% (266) | 54% (263) | 56% (485) | 57% (253) | 54% (232) | 57% (439) | 58% (225) | 56% (214) |
| **Race** | Native American | 2.3% (22) | 2.8% (13) | 1.9% (9) | 2.0% (17) | 2.3% (10) | 1.6% (7) | 2.2% (17) | 2.6% (10) | 1.8% (7) |
| | Asian | 3.4% (32) | 3.2% (15) | 3.5% (17) | 3.4% (30) | 3.6% (16) | 3.2% (14) | 4.1% (32) | 4.4% (17) | 3.9% (15) |
| | Native. Haw. or Pac. Isl. | 0.7% (7) | 0.4% (2) | 1.0% (5) | 0.8% (7) | 0.5% (2) | 1.2% (5) | 0.8% (6) | 0.5% (2) | 1.0% (4) |
| | Black, African American | 6.0% (57) | 5.8% (27) | 6.2% (30) | 5.9% (51) | 6.1% (27) | 5.6% (24) | 5.9% (46) | 6.7% (26) | 5.2% (20) |
| | White | 85% (804) | 85% (394) | 85% (410) | 86% (748) | 86% (379) | 86% (369) | 85% (662) | 86% (335) | 85% (327) |
| | Other/Declined | 4.4% (42) | 3.9% (18) | 5.0% (24) | 4.1% (36) | 3.6% (16) | 4.6% (20) | 4.0% (31) | 2.8% (11) | 5.2% (20) |
| | Race Missing | 0.7% (7) | 0.6% (3) | 0.8% (4) | 0.6% (5) | 0.2% (1) | 0.9% (4) | 0.4% (3) | 0% (0) | 0.8% (3) |
| **Hispanic** | | 12% (112) | 13% (59) | 11% (53) | 12% (107) | 13% (59) | 11% (48) | 11% (86) | 10% (39) | 12% (47) |
| Unknown | | 0.6% (6) | 0.9% (4) | 0.4% (2) | 0.6% (5) | 0.7% (3) | 0.5% (2) | 0.3% (2) | 0.3% (1) | 0.3% (1) |
| **Days since last vaccine** | | 194 (131, 240) | 195 (131, 233) | 193 (132,250) | 195(132, 241) | 196 (131, 234) | 192 (133, 250) | 194 (127,239) | 194(124, 234) | 194(132, 241) |
| **Vaccination status at baseline** | No Vaccine | 45% (428) | 48% (222) | 43% (206) | 44% (384) | 48% (209) | 41% (175) | 43% (334) | 45% (177) | 41% (157) |
| | Primary Series | 50% (471) | 47% (218) | 52% (253) | 51% (440) | 47% (208) | 54% (232) | 51% (397) | 49% (190) | 54% (207) |
| | Booster | 4.9% (46) | 4.8% (22) | 5.0% (24) | 5.4% (47) | 5.2% (23) | 5.6% (24) | 5.7% (44) | 5.9% (23) | 5.5% (21) |
| **Medical History** | BMI | 30 (27, 34) | 30 (27, 35) | 30 (27, 34) | 30 (27, 34) | 30 (27, 35) | 30 (27, 34) | 30 (27, 34) | 30 (27, 35) | 30 (27, 40) |
| | BMI >= 30kg/m2 | 50% (469) | 50% (232) | 49% (237) | 49% (430) | 51% (226) | 47% (204) | 50% (386) | 52% (202) | 48% (184) |
| | Cardiovascular Dis. | 29% (271) | 28% (129) | 29% (142) | 29% (253) | 29% (127) | 29% (126) | 30% (234) | 31% (121) | 29% (113) |
| | Diabetes | 2.1% (20) | 2.8% (13) | 1.4% (7) | 2.1% (18) | 2.7% (12) | 1.4% (6) | 2.2% (17) | 3.1% (12) | 1.3% (5) |
| **Symptom duration (days)** | | 4.7 (1.9) | 4.7 (1.8) | 4.8 (1.9) | 4.7 (1.8) | 4.7 (1.8) | 4.7 (1.9) | 4.8 (1.9) | 4.7 (1.8) | 4.8 (2.0) |
| **<= 4 days of Symptoms** | | 47% (434) | 49% (221) | 45% (213) | 46% (391) | 47% (202) | 45% (189) | 46% (347) | 48% (182) | 44% (165) |
| **Variant Period** | Alpha | 13% (123) | 13% (60) | 13% (63) | 13% (111) | 12% (55) | 13% (56) | 14% (106) | 14% (54) | 14% (52) |
| | Delta | 65% (610) | 64% (297) | 65% (313) | 65% (566) | 66% (289) | 64% (277) | 65% (500) | 65% (252) | 64% (248) |
| | Omicron | 22% (212) | 23% (105) | 22% (107) | 22% (194) | 22% (96) | 23% (98) | 22% (169) | 22% (84) | 22% (85) |
| **Insurance Type** | Private | 66% (619) | 65% (299) | 66% (320) | 66% (572) | 64% (283) | 67% (289) | 67% (520) | 67% (261) | 67% (259) |
| | Medicare | 7.8% (74) | 7.4% (34) | 8.3% (40) | 7.2% (63) | 7.0% (31) | 7.4% (32) | 7.6% (59) | 7.4% (29) | 7.8% (30) |
| | Medicaid | 13% (123) | 14% (63) | 12% (60) | 14% (118) | 14% (62) | 13% (56) | 13% (103) | 13% (51) | 14% (52) |
| | No insurance | 12% (117) | 13% (58) | 12% (59) | 12% (106) | 13% (57) | 11% (49) | 11% (86) | 11% (43) | 11% (43) |
| | Unknown | 1.3% (12) | 1.7% (8) | 0.8% (4) | 1.4% (12) | 1.6% (7) | 1.2% (5) | 0.9% (7) | 1.5% (6) | 0.3% (1) |

[1]Median (IQR); %(N); Mean (SD) . Abbreviations: BMI=body mass index; Dis.=disease

Back to Top

**Table S7. Percentage of viral load specimens undetectable, among observed data**

| Study Day | Fluvoxamine 50mg twice per day | | Ivermectin 430 mcg/kg/d | | Metformin | |
|---|---|---|---|---|---|---|
| | Active | Placebo | Active | Placebo | Active | Placebo |
| Day 1 | 20.7% | 18.8% | 16.1% | 20.9% | 18.0% | 18.0% |
| Day 5 | 48.8% | 45.5% | 46.8% | 51.0% | 50.1% | 45.5% |
| Day 10 | 82.1% | 82.5% | 83.4% | 84.8% | 85.7% | 77.4% |

**Table S8. Mean change in vial load from baseline to follow-up with the primary analytic model, dropping covariates one at a time.**

| Analysis | Effect Type | Effect | SE | 95% CI | p−value |
|---|---|---|---|---|---|
| Unadjusted | Average Effect | −0.656 | 0.285 | (−1.215, −0.097) | 0.021 |
| | Day 5 Effect | −0.489 | 0.280 | (−1.038, 0.06) | 0.081 |
| | Day 10 Effect | −0.835 | 0.429 | (−1.676, 0.006) | 0.052 |
| w/o Lab | Average Effect | −0.562 | 0.254 | (−1.061, −0.063) | 0.027 |
| | Day 5 Effect | −0.481 | 0.238 | (−0.946, −0.015) | 0.043 |
| | Day 10 Effect | −0.650 | 0.400 | (−1.434, 0.134) | 0.104 |
| w/o Other Drugs | Average Effect | −0.553 | 0.252 | (−1.047, −0.059) | 0.028 |
| | Day 5 Effect | −0.471 | 0.235 | (−0.933, −0.01) | 0.045 |
| | Day 10 Effect | −0.641 | 0.395 | (−1.415, 0.134) | 0.105 |
| w/o Baseline VL | Average Effect | −0.511 | 0.267 | (−1.035, 0.013) | 0.056 |
| | Day 5 Effect | −0.360 | 0.265 | (−0.879, 0.16) | 0.175 |
| | Day 10 Effect | −0.674 | 0.395 | (−1.449, 0.101) | 0.088 |
| w/o Vaccination Status | Average Effect | −0.607 | 0.253 | (−1.103, −0.111) | 0.016 |
| | Day 5 Effect | −0.521 | 0.236 | (−0.984, −0.058) | 0.027 |
| | Day 10 Effect | −0.700 | 0.396 | (−1.476, 0.076) | 0.077 |

Back to Top

**Figure S2. Change in viral load from baseline to follow-up, complete case analysis.**

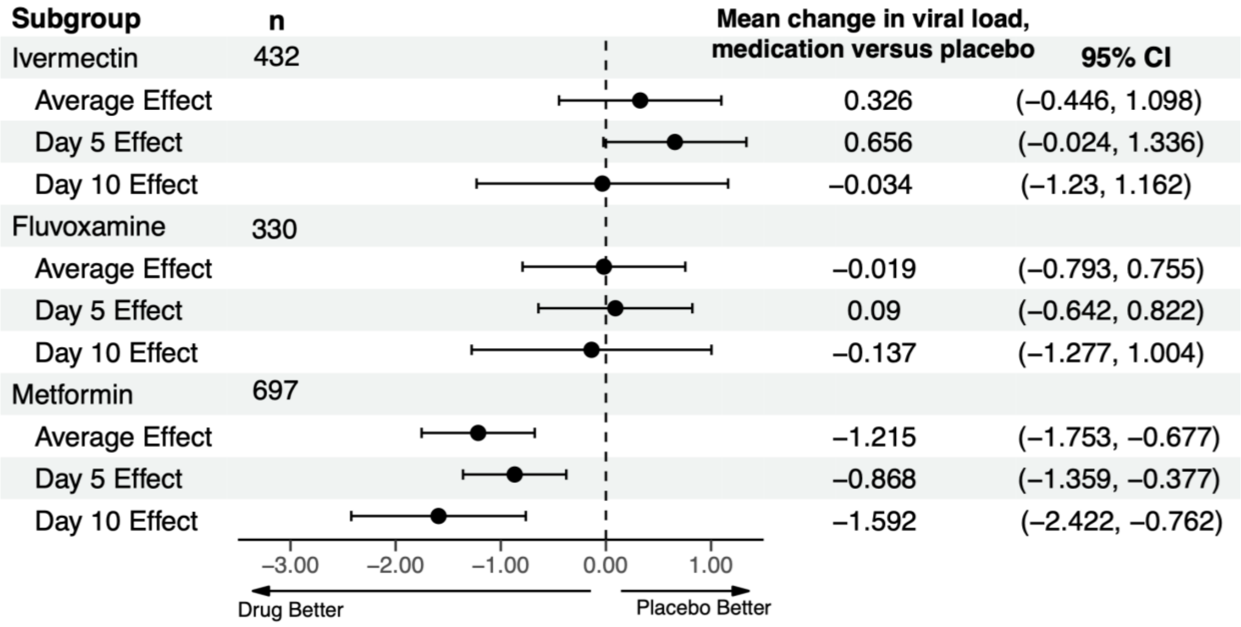


The mean change in SARS-CoV-2 viral load in log10 copies per ml for medication compared to placebo using the primary analytic model, multiply imputed Tobit analysis, in the complete case sample (those who submitted all three samples). The 95% CI denotes a 95% confidence interval. The vertical line indicates the value for a null effect.

Back to Top

**Figure S3. Percent of participants with a detectable viral load at Days 1, 5, 10; the percent of participants with a higher viral load at Day 10 than Day 5, in the complete case sample.**

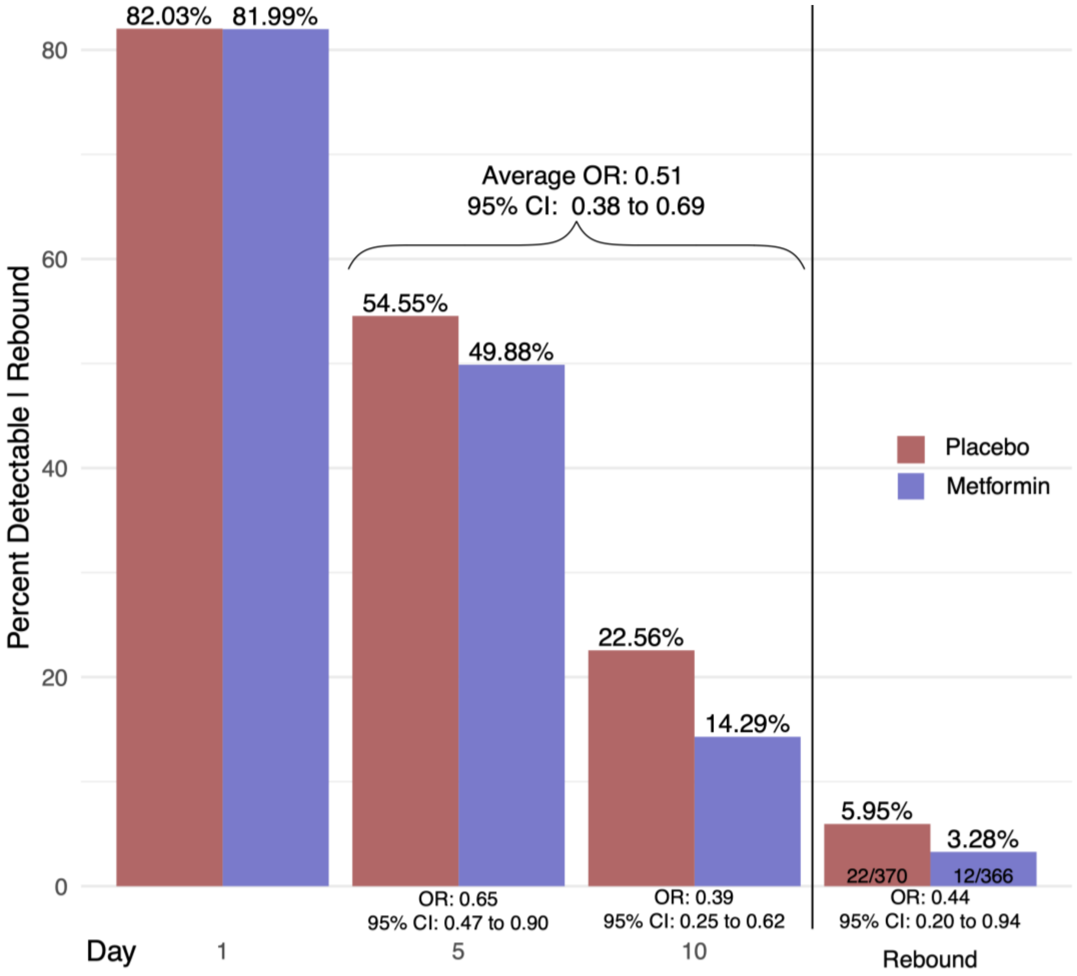


The bar graph depicts the adjusted percent of viral load samples that were detectable at Day 1, 5, and 10. Estimates were based on the adjusted logistic GEE analysis. Odds ratios correspond to adjusted effects on the odds ratio scale. The bars to the right of the vertical line represent the adjusted percent whose day 10 was greater than the day 5 viral load. These analyses were done in the complete case sample (those who submitted all three samples).

Back to Top

**Figure S4. Effect of metformin versus placebo on viral load over time with model prediction, in the complete case sample.**

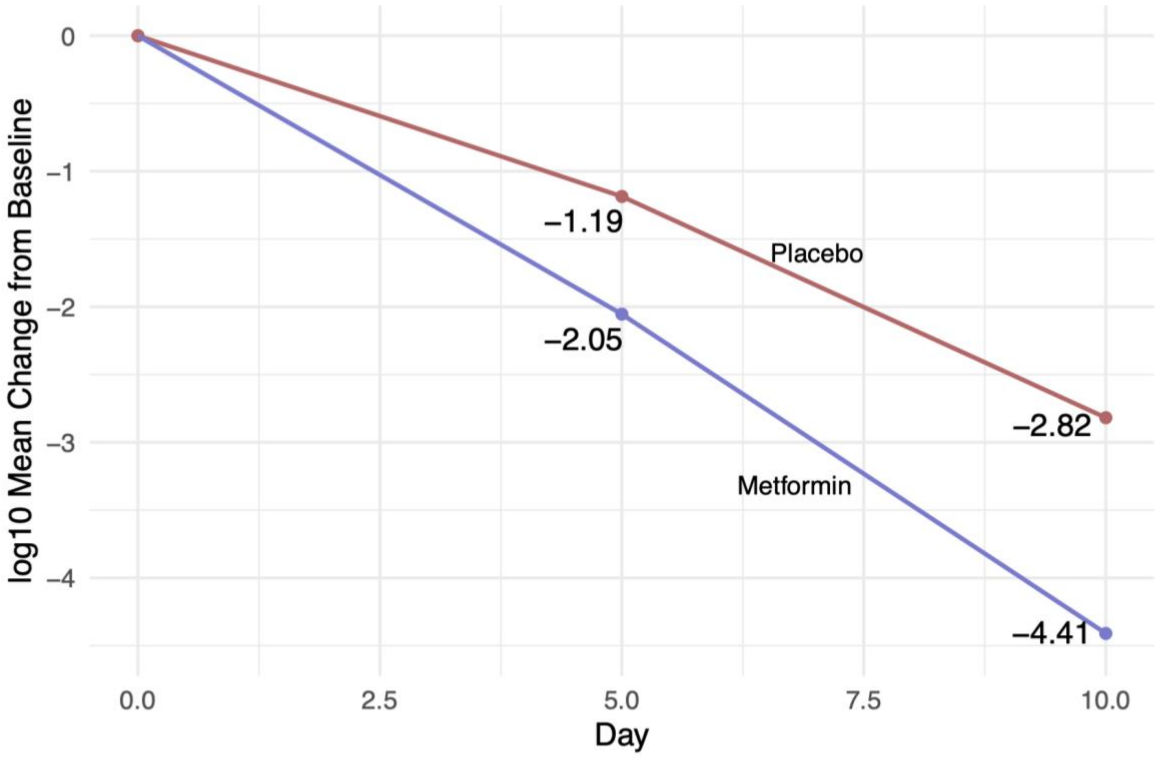


This is a line graph that depicts the adjusted mean change in $\log_{10}$ copies per ml (viral load) from baseline (Day 1) to Day 5 and Day 10. The mean change estimates are based on the primary analytic model, multiply imputed Tobit analysis for undetectable viral loads, in the complete case sample (participants who submitted all three samples).

Back to Top

| **Table S9. Additional detail on analytic approach** |
|---|
| The Gaussian assumption of the Tobit model was assessed by conducting the same analysis with a semiparametric rank-based censored linear regression model that omits the assumption of Gaussian distribution. Results indicated little impact of the Gaussian assumption.<br><br>The exact date and time of specimen collection were utilized for assessing the change over time. The protocol allowed collection of samples to occur +/- 3 days from Days 1, 5, 10, and two thirds were collected within 1 day of those study days. When time of day was not recorded, time was imputed as the courier pickup time (most conservative estimate).<br><br>*Sample Size and Randomization*<br>The sample size for the trial was determined based on the primary clinical outcome. The randomization was stratified by study site and schedules were pre-generated using the mass-weighted urn design which limits deviations from the targeted equal allocation similar to permuted blocks.[a,b] |

| **Table S10. Laboratory Procedures** |
|---|
| Two different labs received and processed samples over the course of the study: the Advanced Research and Diagnostic Laboratory (ARDL) and the University of Minnesota Genomic Center (UMGC). The ARDL laboratory processed samples for the first half of the study period, and the UMGC processed samples for the second half. For each sampled individual, all three time points were processed in the same laboratory. As two different labs processed specimens using different assays and qPCR instrumentation, correction factors were calculated for within-lab and between-lab variation, available upon request.<br><br>ARDL Protocol: Testing at ARDL was performed with the HDPCR SARS-CoV-2 assay (ChromaCode, Carlsbad, CA) on the QuantStudio 7 (Applied Biosystems, Waltham, MA), using a validated extractionless protocol, and results were captured with the StarLims Laboratory Information Management System (Abbott, Chicago, IL). When N1 or N2 were undetected, the cycle threshold value was set to 45, which was the maximum number of PCR cycles. The level of quantification was 80 copies/mL and level of detection was 8 copies/mL.<br><br>UMGC Protocol: Testing at UMGC was performed using a lab-developed solid-phase extraction and qPCR (N1, RNase P) assay on the QuantStudio 5 (Applied Biosystems, Waltham, MA), and results were captured with a proprietary LIMS system. The level of quantification was 1 copy/mL of SARS-CoV-2 virus, which was also the level of detection.<br><br>The differences are primarily related to the input volume of the assay as the ARDL assay utilized 1/10th of the amount of material relative to UGMC, so this smaller volume is equilibrated by using 1/10th of the threshold of quantification for imputation in the primary model. |


a. Zhao W. Mass weighted urn design--A new randomization algorithm for unequal allocations. *Contemp Clin Trials*. Jul 2015;43:209-16. doi:10.1016/j.cct.2015.06.008
b. Bramante CT, Huling JD, Tignanelli CJ, et al. Randomized Trial of Metformin, Ivermectin, and Fluvoxamine for Covid-19. *The New England Journal of Medicine*. Aug 18 2022;387(7):599-610. doi:10.1056/NEJMoa2201662


Back to Top

**Figure S5. Overall results for metformin, separated by the two laboratories.**

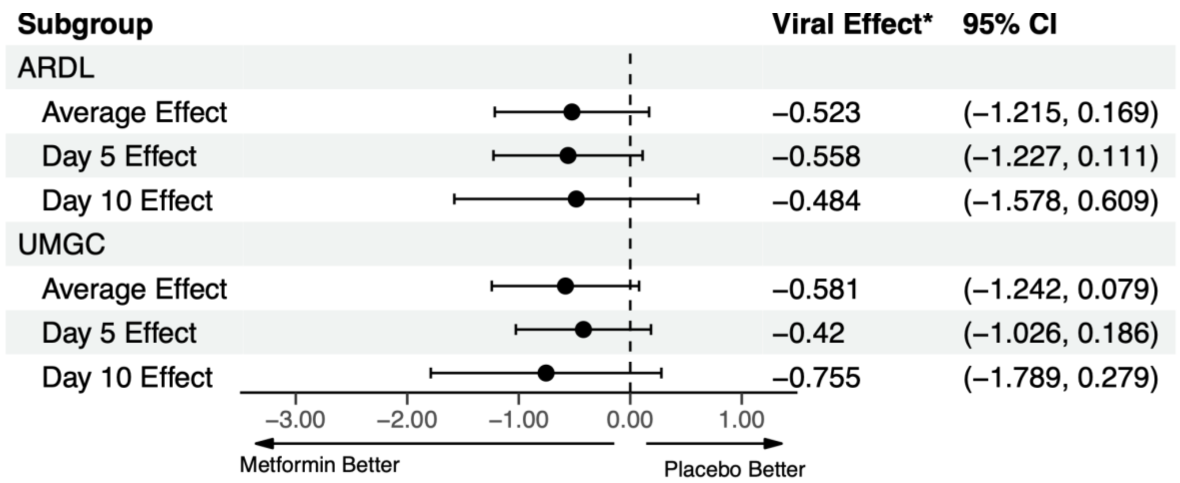


This is a forest plot that depicts the effect of metformin compared to placebo on $\log_{10}$ copies per ml (viral load), overall and at Days 5 and 10. "Viral Effect*" denotes mean change in viral load in $\log_{10}$ copies per ml using the primary analytic model, multiply imputed Tobit analysis for missing and undetectable viral loads. Confidence intervals are 95% confidence intervals. The vertical dashed line indicates the value for a null effect. The top three rows show results for the first lab, the next three rows show results for the second lab.

Back to Top

**Figure S6. Effect of metformin versus placebo on viral load over time using manual midpoint substitution for undetectable viral loads as a sensitivity analysis.**

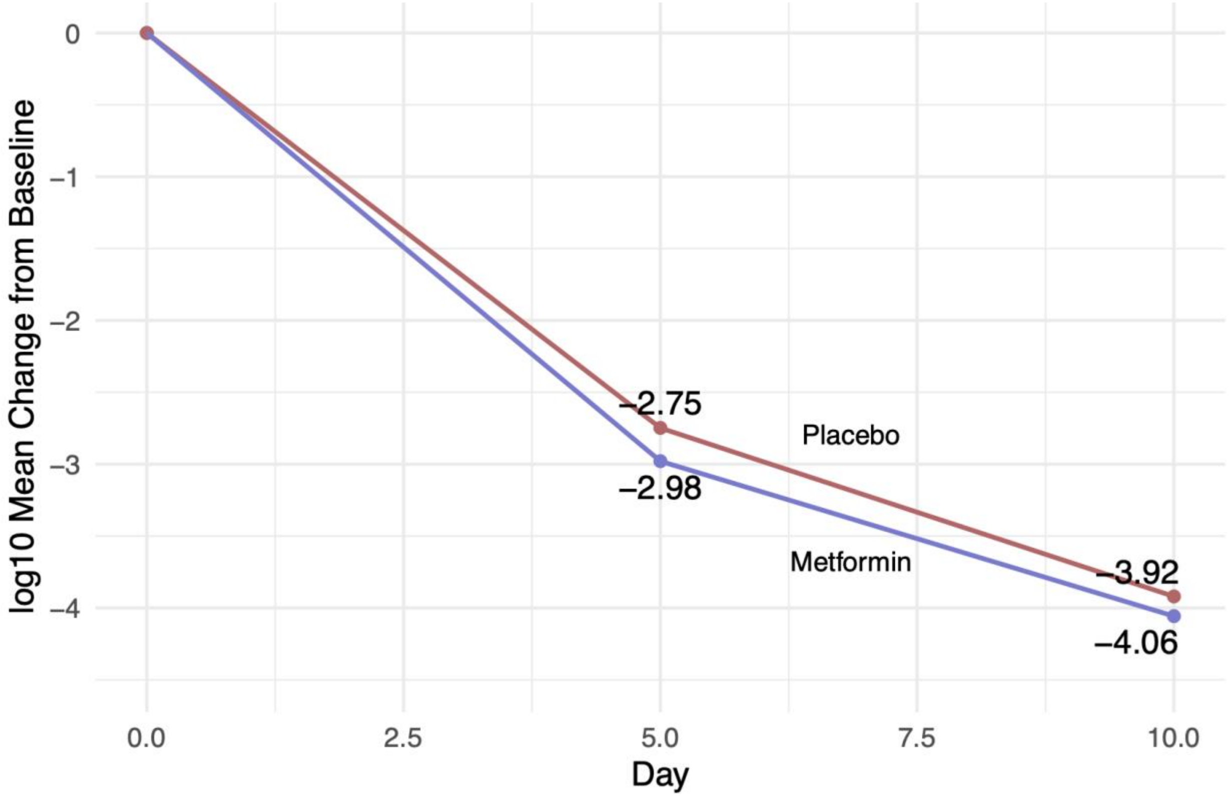


This is a line graph that depicts the adjusted mean change in $log_{10}$ copies per ml (viral load) from baseline to Day 5 and Day 10. The blue line is the metformin group and the red line is the placebo group. The mean change estimates are based on the adjustment variables in the primary analytic model, but where undetectable (i.e. left-censored) viral loads are substituted for values equal to one-half the limit of detection of the lab assay used. The substituted values are then treated as the true viral load value in the analysis.

Back to Top

**Figure S7. Effect of metformin versus placebo on viral load over time using the limit of quantification for imputation for undetectable viral loads, as a sensitivity analysis**.

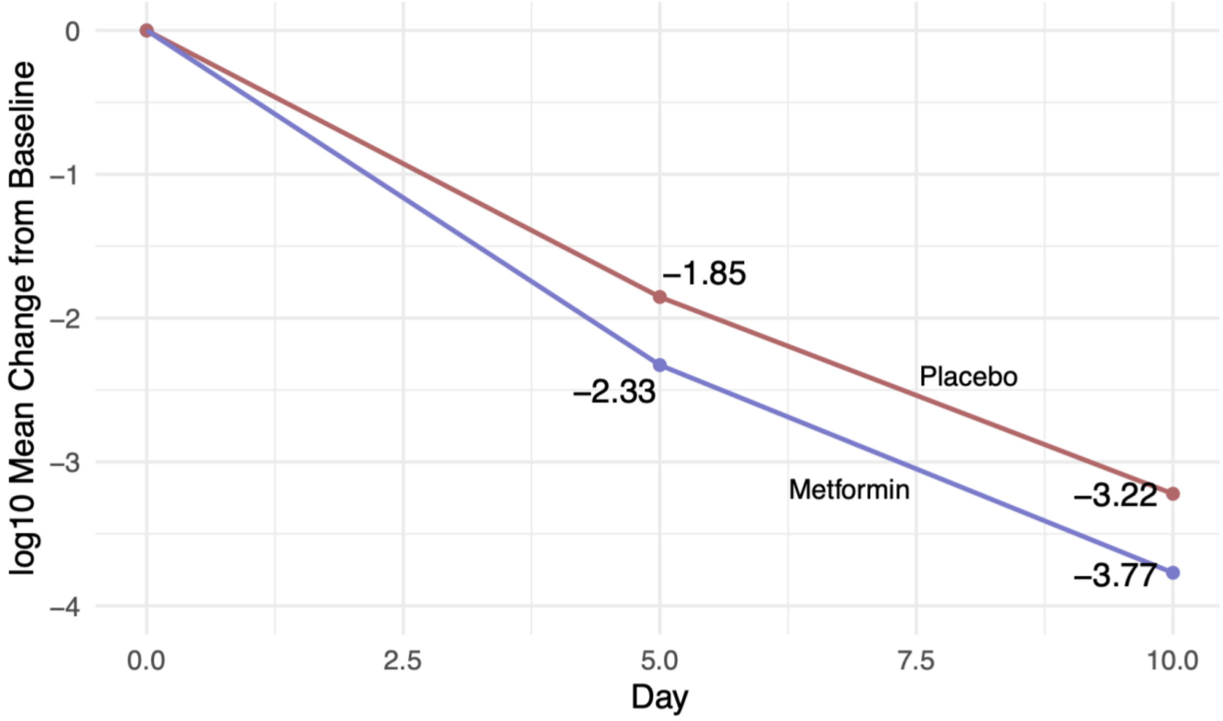


This is a line graph that depicts the adjusted mean change in $log_{10}$ copies per ml (viral load) from baseline (Day 1) to Day 5 and Day 10. The blue line is the metformin group and the red line is the placebo group. The mean change estimates are based on the primary analytic model, multiply imputed Tobit analysis, but with imputing undetectable values from the level of quantification rather than from the level of detection.

Back to Top

**Table S11. Sensitivity analysis excluding patients who obtained their first sample after taking the first 500mg dose of metformin, primary analytic model.**

| Effect Type | Effect | SE | 95% CI | p-value | n |
|---|---|---|---|---|---|
| Exclusion of n=126 sampling ≥12 hours after their first dose | | | | | |
| Average Effect | -0.491 | 0.274 | (-1.029, 0.047) | 0.074 | 873 |
| Day 5 Effect | -0.343 | 0.261 | (-0.855, 0.168) | 0.188 | 873 |
| Day 10 Effect | -0.641 | 0.429 | (-1.481, 0.200) | 0.135 | 873 |
| Exclusion of n=53 sampling ≥24 hours after their first dose | | | | | |
| Average Effect | -0.565 | 0.259 | (-1.07, -0.056) | 0.030 | 946 |
| Day 5 Effect | -0.473 | 0.245 | (-0.95, 0.008) | 0.054 | 946 |
| Day 10 Effect | -0.662 | 0.407 | (-1.459, 0.136) | 0.104 | 946 |

With inclusion or exclusion of the exact time of baseline sampling being before or after the initial 500mg day 1 metformin dosing, the viral effect results are consistent.

Back to Top
